# Bifrontal transcranial direct current stimulation normalises learning rate adjustment in low mood

**DOI:** 10.1101/2023.04.24.23289064

**Authors:** Verena Sarrazin, Margot Overman, Luca Mezossy-Dona, Michael Browning, Jacinta O’Shea

## Abstract

**Background:** Transcranial direct current stimulation (tDCS) applied to dorsolateral prefrontal cortex has mild to moderate antidepressant effects. Little is known about the mechanisms of action. Other antidepressant treatments have been shown to act in part by reducing negative biases, which are thought to play a causal role in the maintenance of depression. Negative biases are hypothesized to stem from aberrant reinforcement learning processes, more precisely from overestimation of the informativeness of negative outcomes. The aim of this study was to test whether bifrontal tDCS might normalise such aberrant reinforcement learning processes in depressed mood.

**Methods:** 85 community volunteers with low mood received tDCS during (or before) the performance of a reinforcement learning task that manipulated the informativeness (volatility) of positive and negative outcomes. In two sessions participants received real or sham tDCS in counter-balanced order. Baseline performance (sham tDCS) was compared to a sample of healthy individuals (n = 40) to identify the effect of low mood on task performance. The impact of tDCS on task performance was assessed by contrasting real and sham tDCS.

**Results:** Low mood was characterised by decreased adjustment of loss relative to win learning rates in response to changes in informativeness. Bifrontal tDCS applied during task performance normalised this deficit by increasing the adjustment of loss learning rates to informativeness. Bifrontal tDCS applied before task performance had no effect indicating that the stimulation effect is cognitive state dependent.

**Conclusions:** Our study provides preliminary evidence that bifrontal tDCS can normalise aberrant reinforcement learning processes in low mood. Crucially, this was only the case if stimulation was applied during task performance, suggesting that combining tDCS with a concurrent cognitive manipulation might increase the functional impact on cognitive functions and potentially on emotional symptoms. Future studies are needed to test if the effect on learning processes might have a beneficial effect on mood itself.

## 1 Introduction

TDCS is a non-invasive brain stimulation method which applies constant electric currents through the scalp to change cortical excitability. Transcranial direct current stimulation (tDCS) is under investigation as an antidepressant treatment. Depression is associated with hypoactivity in the left, and hyperactivity in the right dorsolateral prefrontal cortex (DLPFC)(1-3). In depression trials, a bifrontal tDCS montage is commonly used which applies anodal (excitatory) tDCS to left, and cathodal (inhibitory) tDCS to right DLPFC, with the aim to re-balance activity between these regions (4, 5). A recent meta-analysis suggests that tDCS applied to DLPFC has mild to moderate antidepressant effects (6). More research is needed to understand the mechanisms of action and develop approaches to improving the application of tDCS for depression treatment.

Depression is typically characterised by a negative cognitive bias, i.e. information processing is biased towards negative rather than positive information (2). Compared to healthy controls, individuals with depressive symptoms remember more negative words (7), perceive feedback as more negative (8) and tend to interpret ambiguous information as negative (9). Negative biases are hypothesised to play a causal role in the development and maintenance of depressive symptoms (2, 10, 11). Reduction in negative bias has been shown to be one mechanism of action of antidepressant drugs (10-12). Negative biases are associated with hypoactivity in the left, and hyperactivity in the right DLPFC (1, 3). Bilateral tDCS is applied with the physiological aim to rebalance activity between these areas. This suggests the hypothesis that tDCS might also reduce negative biases.

Recent research in computational psychiatry has shed light on how negative biases might develop. Information processing should prioritise outcomes that are most informative, i.e. most useful for predicting future outcomes (13). How informative outcomes are depends in part on the volatility of the underlying reward association (13, 14). If the association is volatile (i.e. changes over time) compared to stable, an unexpected outcome is more likely to signal a change in the underlying reward association, i.e. it is more informative. In a volatile environment, behaviour should therefore be changed more quickly in response to unexpected outcomes than in stable environments, i.e. learning rates should be higher in volatile environments (13, 14). Learning rates can therefore be interpreted as a measure for estimated informativeness of outcomes.

While human participants adjust their learning rates to volatility (13, 15), anxiety and depression have been associated with deficits in doing so (16, 17). Difficulties tracking informativeness of outcomes might lead to a negative bias if an individual estimates negative events to be more informative than positive events (18, 19). Consequently, an individual would focus their attention on negative rather than positive outcomes. On a computational level, this might manifest in increased punishment vs. reward learning rates (20-22). Increased punishment learning rates might lead to negative behavioural consequences, by causing an individual to give up quickly after receiving negative feedback which might prevent them from experiencing potential future positive outcomes. A potential therapeutic target would therefore be to normalise reward and punishment learning in depression.

The aim of this study was to investigate whether bifrontal tDCS might normalise alterations in reinforcement learning associated with depression. DLPFC is part of a brain network involved in reinforcement learning and has been found to be activated in response to volatility (23-25). In our previous study, we found that bifrontal tDCS increased reward learning rates in healthy volunteers (26). However, due to the alterations in reinforcement learning in depression, it is unclear whether the same tDCS effect should be expected in individuals with low mood. To assess this, we compared task performance between healthy volunteers from our previous study and individuals with depressive symptoms (26). We hypothesised that individuals with depressive symptoms would show increased punishment vs. reward learning rates (20-22) and/or reduced adjustment of learning rates to volatility (16, 17). We then tested whether bifrontal tDCS applied during task performance might normalise the observed behavioural alterations in the low mood participants.

Our secondary aim was to test the hypothesis that tDCS would have a greater functional impact when applied during rather than before task performance. When applied during activity-dependent neuroplasticity (‘online’) tDCS has been shown to change brain and behaviour, with no such effect when stimulation was instead applied ‘offline’ prior to plasticity induction (26-29). We therefore hypothesised that tDCS would normalise alterations in reinforcement learning only if applied *during*, but not *before* task performance.

## 2 Methods and Materials

This study has been pre-registered (https://clinicaltrials.gov/ct2/show/NCT03393312).

### 2.1 Sample

85 participants suffering from low mood (Beck Depression Inventory II (30) score of at least 10) were recruited via university email lists and social media advertisement (see Table 1 for demographic details). 41 participants were assigned to the “tDCS during task” group, and 44 participants to the “tDCS before task” group. All participants completed two testing session in which they received real or sham tDCS in counter-balanced order. An a-priori power analysis based on the effect size from our previous study in healthy participants (26) indicated that a sample size of 38 participants per group was required to reach a power level of 80% for contrasting the effect of real vs. sham tDCS (paired t-test, two-tailed, *Cohen’s dz* = 0.472). This study was approved by the University of Oxford Central University Ethics Committee (R67041/RE002). All participants gave informed written consent to take part in the study.

**Table 1.**
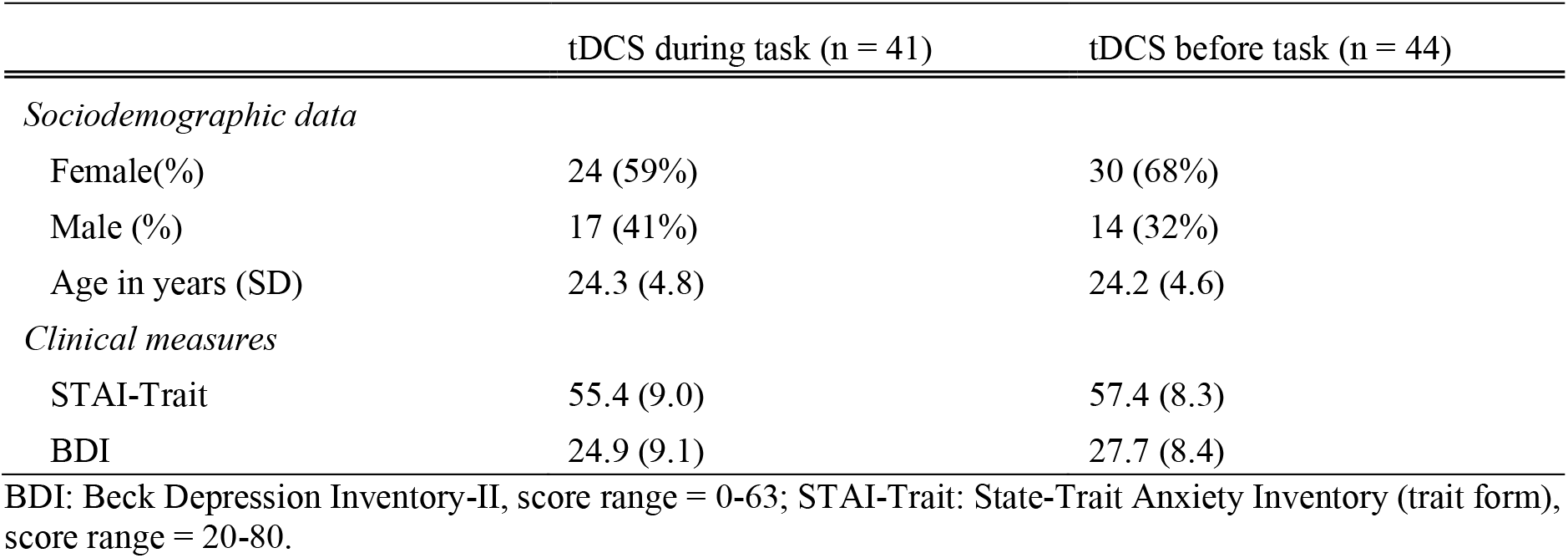
Mean (SD) baseline characteristics for the “tDCS during task” and “tDCS before task” groups.

To investigate the effect of low mood on learning behaviour in our experimental paradigm, we compared task performance during sham tDCS between participants with low mood and the sample of healthy participants from our previous study (26). To avoid confounds with learning effects from task repetition, only individuals (from either study) who received sham tDCS in their first session were included in this analysis (low mood: n = 43, healthy: n = 40). Demographic data and baseline questionnaire scores for both samples are shown in Table 1 and Figure 1.

**Figure 1.**
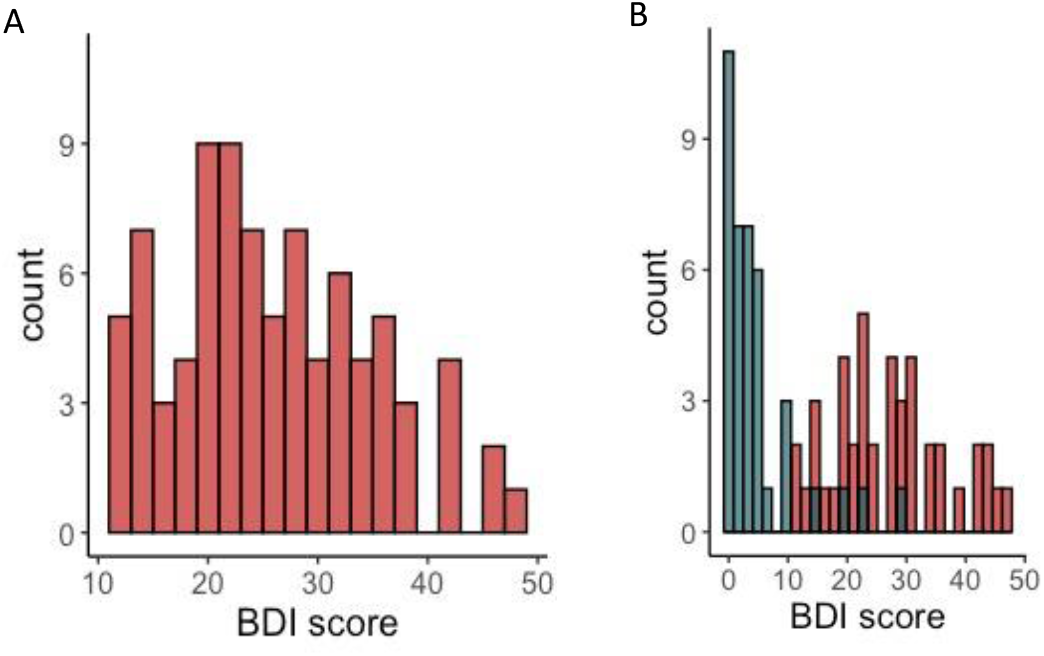
Distribution of BDI scores. (A) Distribution of BDI scores of all participants recruited for this study (n = 85). (B) Comparison of BDI scores for participants included in the comparison between the low mood sample (n=43) and general population sample (n=40). Participants with low mood had significantly higher BDI scores than participants in the general population sample (two-sample Welch t-test: *t*(73.8) = 12, *p* < .001).

### 2.2 Information Bias Learning Task

The Information Bias Learning Task (IBLT)(18) manipulates the relative informativeness of win and loss outcomes. The task is described in detail in Figure 2. In brief, participants performed six blocks of 80 trials in which they were asked to choose between two shapes. Each trial resulted in the receipt of a win (+10p), a loss (−10p), both win and loss (0p), or neither outcome (0p). Wins and losses were independently associated with the two shapes which allowed for separate estimation of win and loss learning rates. The relative informativeness of wins and losses was manipulated throughout the six blocks, such that wins and losses were equally informative (both-volatile), wins were more informative than losses (wins-volatile) or losses were more informative than wins (losses-volatile).

**Figure 2.**
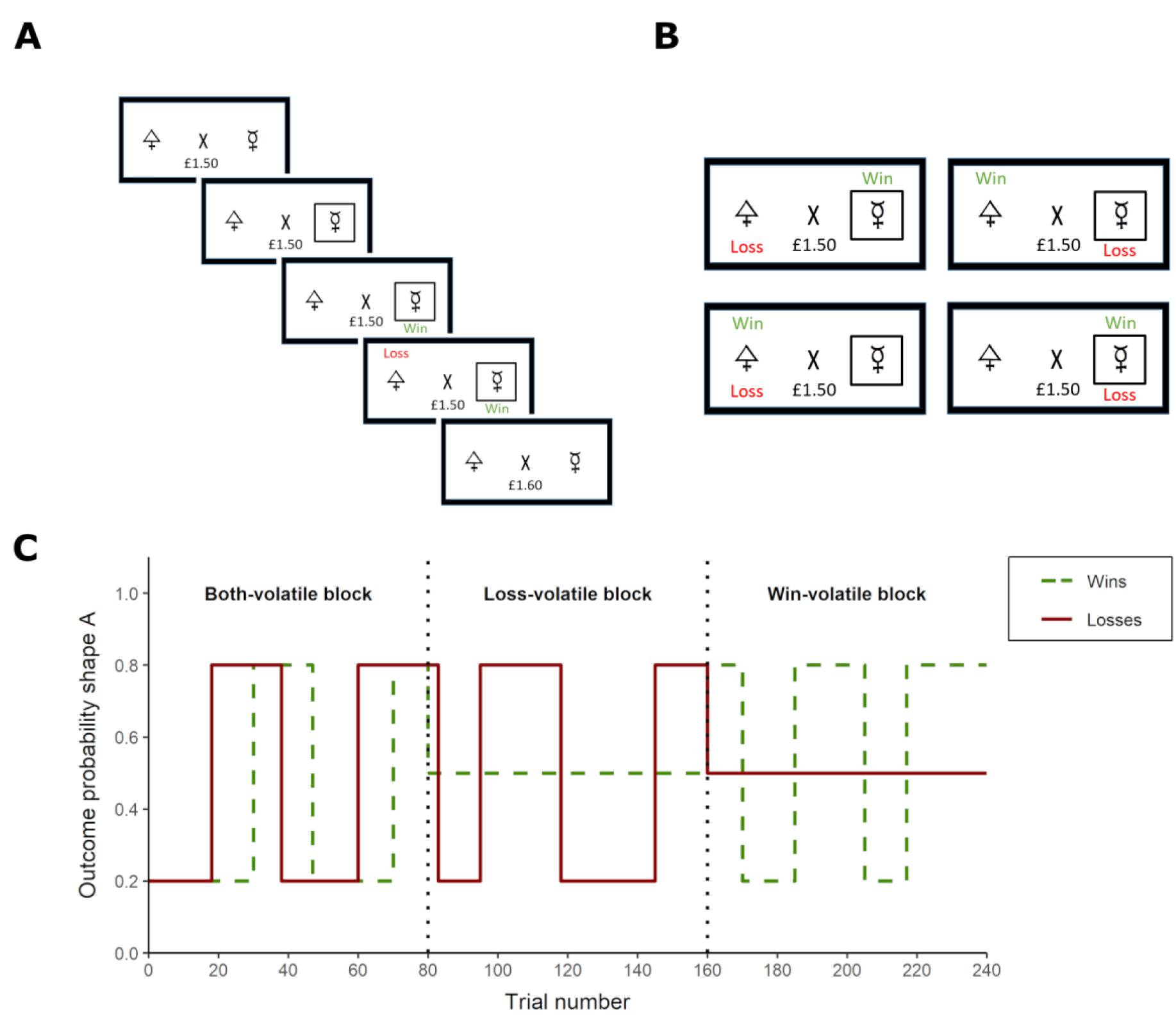
Task design of the Information Bias Learning Task. (A) On each trial, participants were asked to choose one of two shapes, by pressing the keys ‘A’ or ‘L’ for the left or right shape, respectively. Subsequently, a win and a loss outcome appeared on the screen. (B) The win and loss outcomes were independent of each other, resulting in four possible scenarios: The chosen shape might be associated with the win (+10p), the loss (−10p), both outcomes (0p) or neither (0p). Wins and losses were associated with an actual win or loss of 10p on each trial, respectively. (C) Underlying reward structure for a ‘both-volatile’, ‘losses-volatile’ and ‘wins-volatile’ block. In this task, the volatility of the wins and losses was manipulated independently. In ‘wins-volatile’ blocks, the wins were associated with one of the shapes in 80% of the trials, and with the other in 20%. This association reversed a few times within the block. Losses were randomly presented with either shape (50%) and are therefore uninformative. In ‘losses-volatile’ blocks, the probability pattern was reversed. In ‘both-volatile’ blocks, both wins and losses were independently associated with one shape in 80% and with the other in 20. Retrieved from (26).

### 2.3 tDCS protocol

All participants took part in two testing sessions in which they received real or sham tDCS in counter-balanced order (double-blinded). Real tDCS was applied for 20 minutes at an intensity of 2mA. The anode and cathode were placed over left and right DLPFC, respectively, approximated by the F3 and F4 electrode positions (international 10-20 system)(see Figure 3C for a simulation of the electric field). The first 41 participants received tDCS during performance of the second and third task block (“tDCS during task” group, Figure 3A). To test whether the cognitive state during stimulation is critical, another 44 participants received tDCS at rest after the first task block, and performed the remaining task blocks after the stimulation period (“tDCS before task” group, Figure 3B).

**Figure 3.**
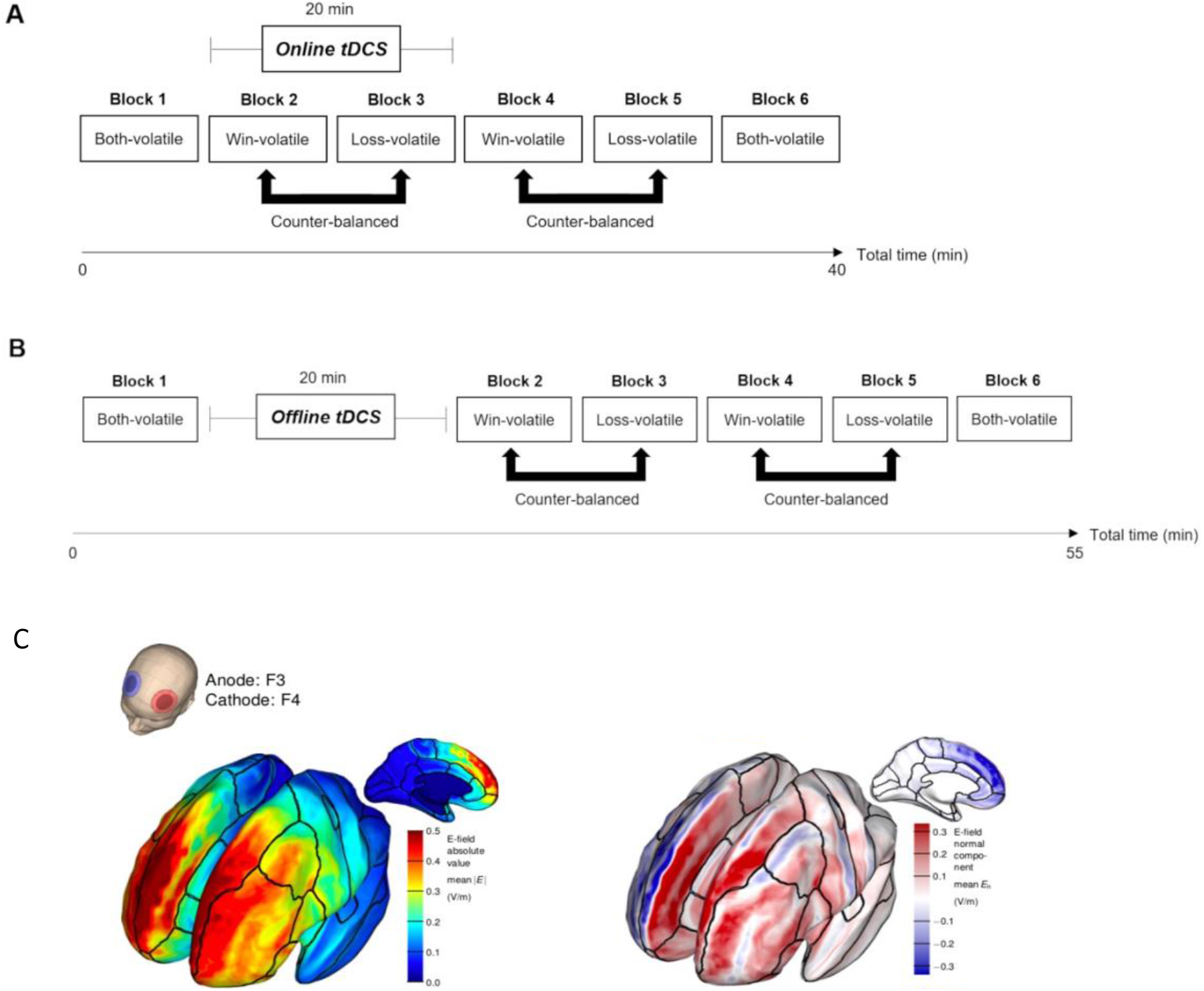
(A) Task protocol for the “tDCS during task” group. Participants started with a ‘both-volatile’ block, and then underwent two ‘wins-volatile’ and two ‘losses-volatile’ block in alternating order. Half of the participants performed the ‘wins-volatile’ block first, while the other half performed the ‘losses-volatile’ block first. The experiment ended with another ‘both-volatile block’. Stimulation was applied during the performance of block 2 and 3. (B) Task protocol for the “tDCS before task” group. The task protocol was identical to (A) with the exception that tDCS was applied at rest after performance of the first task block. (C) Modelling of the electric field induced by the bifrontal tDCS setup, with the anode over left, and the cathode over right DLPFC. The left figure shows the strength of the electric field. The right figure displays the normal component (red = anodal stimulation, blue = cathodal stimulation). Adapted from (26). (C) is adapted from (31) with permission.

### 2.4 Computational modelling

Performance in the IBLT was analysed using computational models that were fit to participants’ trial-by-trial choices. The fit of six models was compared using the Bayesian Information Criterion (BIC) averaged across participants (see Supplementary Material 1). In line with our previous study (26) we first fitted the six models separately to each task block for each participant and session (“block-wise models”). Since the inverse temperature estimates did not vary between task conditions in the original study (18), we applied a second modelling approach in which the inverse temperature parameters were fitted across all six task blocks while the learning rates were allowed to vary between blocks (“constant models”). For each of these modelling approaches, model comparison was performed using six comparator models. Statistical analysis was performed based on the parameter estimates derived from the winning model for each of the two modelling approaches.

The winning block-wise model used a modified version of a Rescorla-Wagner updating rule in which the probability of an outcome being associated with shape A was modelled separately for win and loss outcomes:

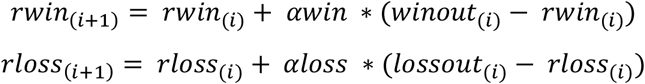

Where *rwin*_(*i*+1)_ and *rloss*_(*i*+1)_ are the estimated probabilities of the win or loss being associated with shape A on trial *i+1*. These probability estimates were updated on each trial with the prediction error on the previous trial weighted by the win or loss learning rate, *αwin* or *αloss*. A Softmax function was used to transform the probability estimates into choice probabilities:

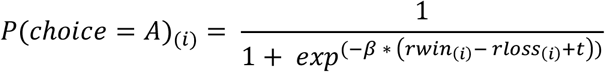

Where *P(choice = A)*_*(i)*_ represents the probability of the participant choosing shape A on trial *i*. The inverse decision temperature *β* captures choice stochasticity (lower values indicate more random behaviour). The tendency parameter *t* accounts for a potential tendency of choosing one shape over the other.

The winning constant model calculated the outcome probabilities as above, but contained two separate inverse temperature parameters for wins and losses (but no tendency parameter):

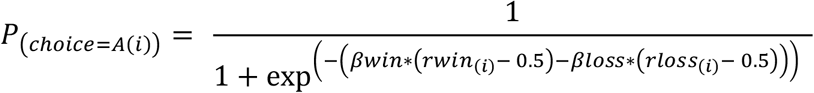

Parameters for the block-wise models were estimated by calculating the posterior distribution over the full parameter space and deriving the expected value of the marginal distribution for each parameter. The constant models were fit to the whole session simultaneously and therefore included a larger number of parameters. Parameters for the constant models were therefore estimated in STAN (32)(Supplementary Material 1.2).

### 2.5 Statistical analysis

The main outcome measures of interest were win and loss learning rates, and the relative adjustment of learning rates between volatile versus stable blocks. *Win learning adjustment* and *loss learning rate adjustment* were defined as:

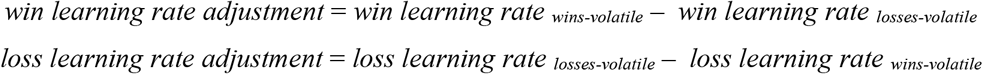

Positive learning rate adjustment values indicate that learning rates were higher in volatile than in stable conditions. To capture the extent to which learning rate adjustment was biased towards either win or loss outcomes, *learning rate adjustment bias* was defined as:

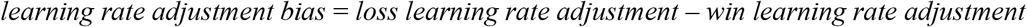

A positive value on *learning rate adjustment bias* therefore indicates that win learning rates were adjusted more to changes in informativeness than loss learning rates. An inverse logit transformation was applied to the learning rate estimates.

As non-computational control analysis, logistic regressions were run to predict the choice on each trial using win and loss outcomes of the previous 3 trials:

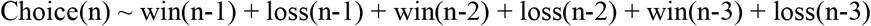

Regressions were run for each task block and each participant individually. To enable comparison across blocks and participants, the regression weights obtained in each regression were divided by the largest weight obtained in that regression so that weights varied between 0 and 1. The analysis of regression weights provides a non-computational alternative to learning rates and is independent of the computational models. Regression weights for more recent outcomes should be larger than regression weights for more distant outcomes. A higher learning rate would correspond to a higher weight on the most recent outcome. The primary measure of interest was therefore the weight for trial n-1.

All analyses were performed in RStudio (Version 1.4.1717, R 4.1.1). All dependent variables were analysed in mixed ANOVAs (ezANOVA package). To test whether low mood might be associated with increased loss vs. win learning rates, learning rate estimates were analysed in ANOVAs including the factors Sample (low mood vs. general population), Valence (win vs. loss), Volatility (both-volatile, wins-volatile and losses-volatile) and Time (first half vs. second half). To test whether low mood might be characterised by decreased learning rate adjustment, an ANOVA including Sample, Valence, and Time tested for a different between the groups on learning rate adjustment. ANOVA including Sample and Time was used to test for a difference in *Learning rate adjustment bias* between the groups.

To test whether tDCS might modulate any of these outcome measures, mixed ANOVAs were run for task performance in blocks 2 and 3 during the stimulation period (or right after the stimulation period for the “tDCS before task” group). For the analysis of learning rates, tDCS Condition (real vs. sham), Valence and Volatility (wins-volatile vs. losses-volatile) were included. For learning rate adjustment, TDCS Condition and Valence were included, for *learning rate adjustment bias*, only tDCS Condition was included. All ANOVAs included Block Order (wins-volatile first vs. losses-volatile first) as between-subject factor of no interest. All analyses were repeated after removing outliers (see Supplementary Material 2-4). Summary statistics for all computational parameters are provided in Supplementary Material 6.

All outcome measures were correlated to BDI and trait anxiety scores. Significance of correlations was assessed using t-tests.

All analyses were repeated after removing outliers. A datapoint was identified as an outlier if it was more than 1.5 times the interquartile range below the first or above the third quartile. For each outcome measure, outliers were removed separately for the levels of the factors of interest (i.e. separately for the general population and low mood sample, and win and loss outcomes where appropriate). For the analysis of the effect of real vs. sham tDCS, a datapoint was identified as outlier if the difference between real minus sham tDCS was more than 1.5 times the interquartile range below the first or above the third quartile. Statistics are reported for the entire dataset unless outlier removal had an impact on the results (for completeness, figures including outliers are included in the supplementary material for all analyses that outlier removal had an impact on).

## 3 Results

### 3.1 Low mood is associated with alterations in learning rate adjustment

In the block-wise model, low mood had no effect on learning rates, learning rate adjustment or *learning rate adjustment bias* (all *p* > .16)(Supplementary Material 3.2).

This contradicts previous findings showing that depression and anxiety are associated with difficulties adjusting learning rates in response to changes in informativeness (16, 17). We therefore tested whether an alternative modelling approach (‘constant model’) would be able to capture this effect in our dataset. In the second modelling approach, the inverse temperature was kept constant across the whole session while the learning rates were allowed to vary between blocks. Changes in choice behaviour between blocks will therefore be attributed to changes in learning rates. There was no effect of Sample on the learning rate estimates derived from the constant model (all p > .07, Figure 4A). However, there was a significant Sample x Valence interaction (*F*(1,72) = 4.4, *p* = .038, seven outliers removed). Post-hoc tests indicated that there was no significant main effect of Sample on win learning rate adjustment (*F*(1,72) = 1.3, *p* = .24) but a trend towards lower loss learning rate adjustment in the sample with low mood (*F*(1,72) = 3.2, *p* = .076)(Figure 4B). In line with the interaction of Sample and Valence on learning rate adjustment, individuals with low mood showed a lower *learning rate adjustment bias* (main effect of Sample: *F*(1,72)= 4.4, *p* = .038, *Cohen’s d* = 0.44)(Figure 4C). That is, while the general population adjusted their win and loss learning rates to a similar extent (*learning rate adjustment bias* not different from zero (one-sample t-test): *t*(35) = -1.3, *p* = 0.17), individuals with low mood adjusted their loss learning rate significantly less than their win learning rate (*learning rate adjustment bias* significantly below zero: *t*(39) = -3.2, *p* = .002). In line with this, there was a negative correlation between BDI score and *learning rate adjustment bias* across groups (r = -.25, *t*(75) = -2.2, *p* = .027; correlations within groups were non-significant, see Supplementary Figure S12).

**Figure 4.**
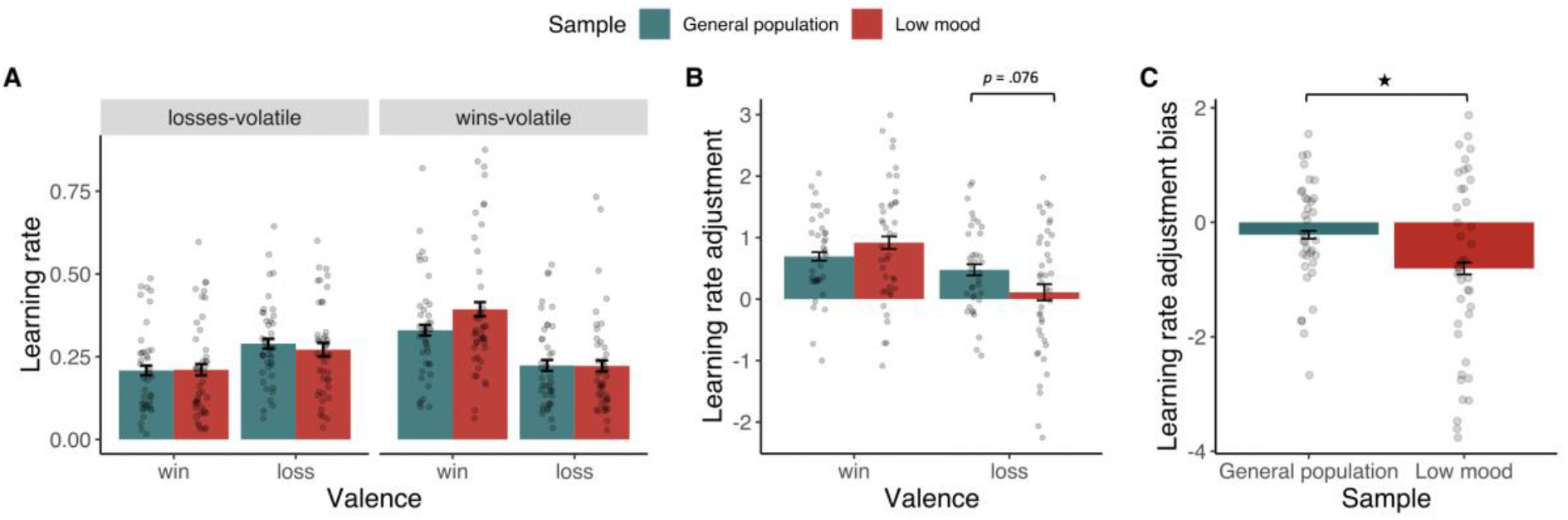
Effect of low mood on learning rate measures. (A) Low mood did not affect learning rates per se. (B) Interaction effect between Sample and Valence on learning rate adjustment. Participants with low mood showed a trend towards lower loss learning rate adjustment. (C) In line with this, participants with low mood showed a significantly lower learning rate adjustment bias.

Previous studies showed that depression and anxiety are associated with decreased adjustment of learning rates to volatility (16, 17). Since the constant model was able to capture this effect in our study, the analysis of the effect of tDCS will focus on the constant model (the tDCS effect in the block-wise model is reported in Supplementary Material 4.3). Additional non-computational analyses were carried out to validate the findings from the constant model (Supplementary Material 5).

### 3.2 Online bifrontal tDCS normalises learning rate adjustment

TDCS had no effect on learning rates per se (all *p* > .60, Figure 5A). However, there was a significant tDCS Condition x Valence interaction on learning rate adjustment (*F*(1,34) = 7.47, *p* = .009, three outliers removed). Real compared to sham tDCS led to an increase in loss learning rate adjustment (*F*(1,34) = 6.0, *p* = .018, *Cohen’s dz* = 0.46), and to a marginally significant decrease in win learning rate adjustment (*F*(1,34) = 4.0, *p* = .051, *Cohen’s dz* = 0.48; Figure 5B). In line with this, real vs. sham tDCS increased *learning rate adjustment bias* (main effect of tDCS: *F*(1,34) = 7.4, *p* = .009, *Cohen’s dz* = 0.65, Figure 5C). During sham tDCS, *learning rate adjustment bias* was negative and significantly different from zero (*t*(37) = 2.1, *p* = .037), indicating that participants adjusted their loss learning rates significantly less than their win learning rate. During real tDCS, there was a trend towards a positive *learning rate adjustment bias*, i.e. towards higher loss than win learning rate adjustment (*t*(37) = 1.8, *p* = .067). The effect on learning rate adjustment did not outlast the stimulation period (no significant effect of tDCS Condition in block 3 and 4, Figure S14) We further hypothesised that the effect of bifrontal tDCS would be specific to the cognitive state during stimulation, i.e. tDCS applied *before* task performance should not have the same effect as tDCS applied *during* task performance. In line with this, tDCS before task performance had no effect on learning rates, learning rate adjustment or inverse temperature (all p > .16). The effect of tDCS during task performance on *learning rate adjustment bias* was marginally though not significantly larger than the effect of tDCS before task performance (*t*(78.9) = -1.5, *p* = .063 (Welch two sample t-test, one-sided))(Figure 6).

**Figure 5.**
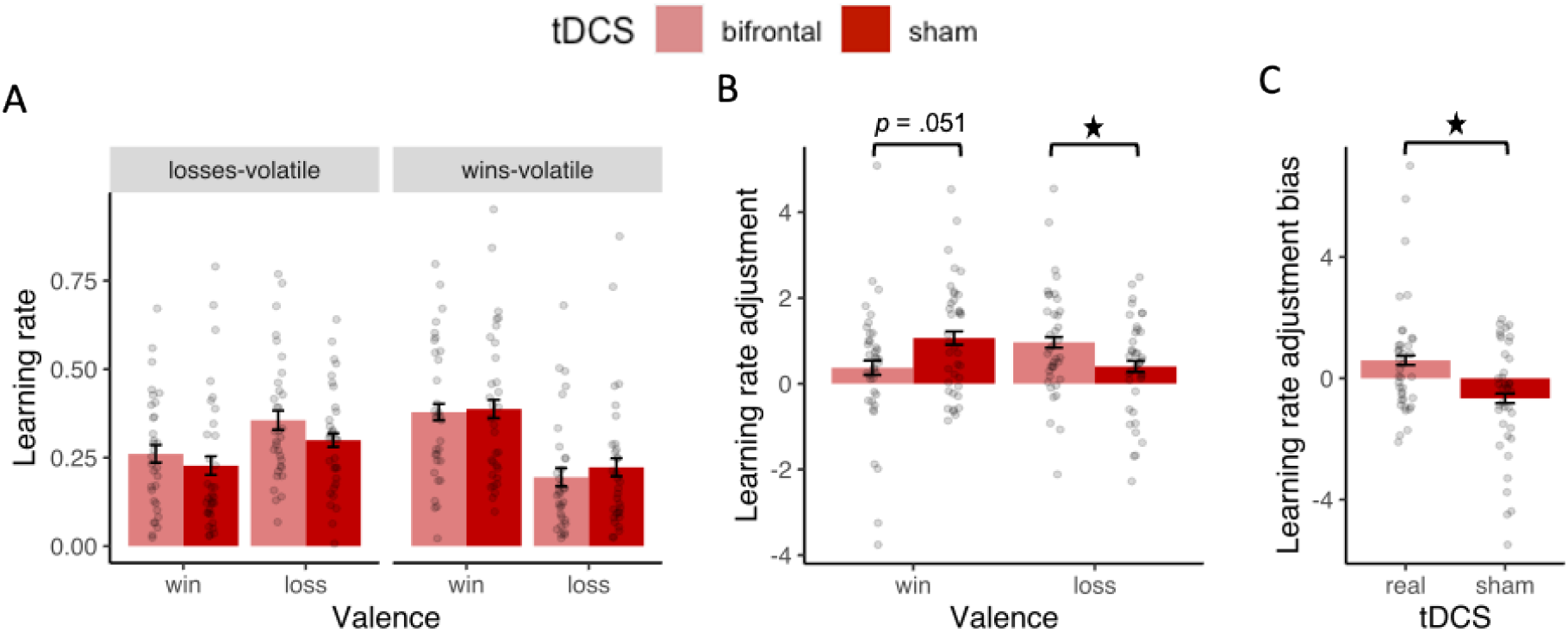
Effect of tDCS applied during task performance on learning rate measures. (A) tDCS had no effect on learning rates per se. (B) TDCS led to significant increase in loss learning rate adjustment, and marginal decrease in win learning rate adjustment. (C) TDCS induced a significant increase in learning rate adjustment bias.

**Figure 6.**
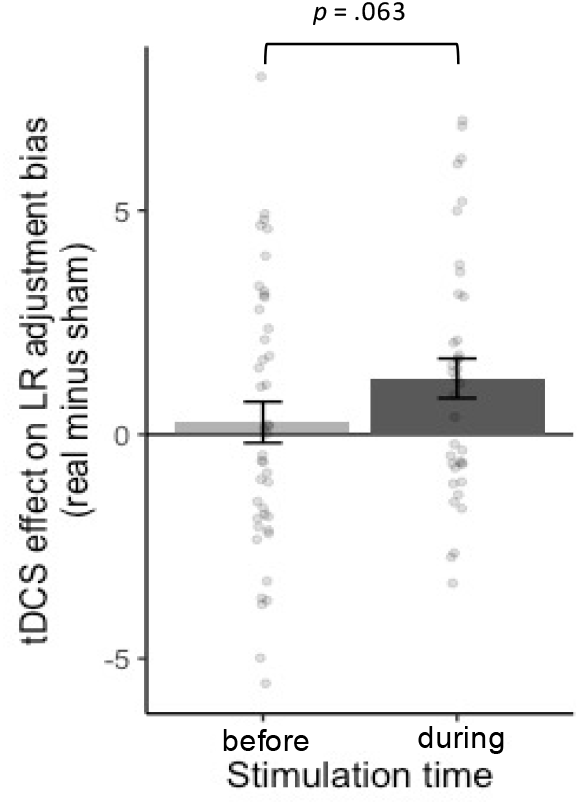
Comparison of the effects of bifrontal tDCS applied *during* or *before* task performance on learning rate adjustment bias. The effect of tDCS applied during task performance was marginally larger than the effect of tDCS applied before task performance.

### 3.3 Non-computational validation

Since the findings obtained from the constant model were not observed in the block-wise model, we conducted control analyses to test whether these findings relate to any non-computational measures. As non-computational analysis, logistic regressions were conducted to predict the trial-wise choices using the win and loss outcomes from the previous three trials (see Supplementary Material 5).

Regression weights for trial-by-trial outcomes capture similar behavioural characteristics as learning rates without relying on a specific computational model. A higher learning rate would correspond to higher weight on the most recent outcomes. During real vs. sham tDCS, individuals with low mood adjusted the weight on the loss outcome from the previous trial more to informativeness than during sham tDCS (marginal effect of tDCS on loss learning rate adjustment: *p* = .069; Figure S21A). This is equivalent to the observed increase in loss learning rate adjustment. Regarding the effect of low mood on learning rate adjustment, the findings from the regression analysis are mixed (Figure S23).

## 4 Discussion

The goal of this study was to investigate whether bifrontal tDCS might normalise alterations in reinforcement learning associated with depression. Participants with low mood performed a task in which the relative informativeness of positive and negative outcomes was manipulated. In comparison to a sample of healthy participants, individuals with low mood did not show increased punishment vs. reward learning rates. However, low mood was associated with reduced adjustment of punishment compared to reward learning rates to changes in informativeness. Bifrontal tDCS applied during task performance increased adjustment of loss compared to win learning rates. This effect was dependent on the cognitive state during stimulation. Bifrontal tDCS applied *before* task performance had no effect. As a limitation of this study, these findings were only observed in one of the two computational models.

Negative biases in depression are hypothesised to arise from alterations in reinforcement learning. Depression has been associated with deficits in adjusting learning rates to the informativeness of outcomes (16, 17). Deficits in evaluating informativeness might lead to negative biases if the informativeness of negative outcomes is overestimated (18, 19) which might be reflected in increased punishment vs. reward learning rates (20-22). In this study, there was no evidence for increased loss vs. win learning rates in individuals with low mood. Several other studies failed to detect an imbalance in reward vs. punishment learning rates in depression (33-35). However, we found that participants with low mood adjusted their learning rates in a different way to changes in informativeness than healthy participants. While healthy participants adjusted their win and loss learning rates to an equal extent, participants with low mood adjusted their loss learning rate less than their win learning rate. This seemed to be caused by both decreased adjustment of loss learning rates, and increased adjustment of win learning rates. Reduced adjustment of loss learning rates to informativeness has previously been associated with depression and anxiety (16, 17). The increase in win learning rates adjustment was unexpected. In contrast to previous studies, our paradigm required simultaneous tracking of rewards and punishments. One potential explanation for the increase in win learning rate adjustment might be that participants with low mood had difficulties tracking the informativeness of losses and therefore focused their cognitive resources on tracking the informativeness of wins as a compensatory strategy.

Bifrontal tDCS applied during task performance normalised learning rate adjustment in individuals with low mood, by increasing the adjustment of loss learning rates, and (marginally) decreasing the adjustment of win learning rates to informativeness. To our knowledge, this is the first evidence suggesting that tDCS might normalise aberrant reinforcement learning processes in individuals suffering from depressive symptoms. While these alterations in reinforcement learning have been suggested to be a potential mechanism leading to negative biases (18, 19), we did not observe a negative bias per se in the task (i.e. no increase in punishment vs. reward learning). Further research is needed to test how alterations in learning rate adjustment relate to negative biases, and whether normalising learning rate adjustment might have beneficial effects in depression treatment.

In our previous study in healthy participants, bifrontal tDCS selectively increased reward learning rates. This raises the question why bifrontal tDCS had a different effect in participants with low mood in this study. We speculate that this might be the case because participants with low mood showed altered learning rate adjustment at baseline and tDCS has been shown to interact with baseline behaviour (36). In individuals with low mood, bifrontal tDCS might normalise aberrant brain activity and thereby normalise information processing. The same tDCS protocol applied to a healthy brain might increase a pre-existing optimism bias which is associated with intact mental health (37, 38).

TDCS affected learning rate adjustment only when applied *during*, but not when applied *before* task performance. This suggests that the cognitive state during the stimulation period is critical. In clinical trials, tDCS is usually applied at rest. However, tDCS applied during activity-dependent plasticity has been shown to potentiate learning effects (27, 39), and might therefore be more effective therapeutically if applied during a learning task relevant to depression. A suitable task might be a training paradigm which trains participants in appropriately adjusting behaviour to changes in informativeness of positive and negative outcomes. A future clinical trial could test whether bifrontal tDCS applied during task performance might be more effective than tDCS applied at rest.

This study was designed to investigate how bifrontal tDCS, a stimulation setup commonly applied in depression trials, affects reinforcement learning in individuals with low mood. Since this setup stimulates large parts of the brain, we can only speculate which neural mechanisms the behavioural effect might rely on. The DLPFC (23, 24) as well as the dorsal anterior cingulate (13) are hypothesised to be involved in adjusting behaviour to volatility. While the bifrontal setup stimulated the DLPFC directly, the dorsal anterior cingulate might be stimulated more indirectly via anatomical connections. Future studies combining tDCS with neuroimaging are needed to investigate how the behavioural effect of tDCS relates to physiological changes in these brain regions.

As a limitation of this study, it should be noted that we used two different computational modelling approaches. The effects on learning rate adjustment were only observed in the model in which the inverse temperature was kept constant across all blocks. It is unclear why different findings were obtained in the block-wise modelling approach which has been applied in previous studies (18, 26, 40). As a control analysis, we ran logistic regressions to test whether non-computational measures might explain the change in learning rate adjustment observed in the constant model. We found that real compared to sham tDCS increased adjustment of the regression weight of the previous loss outcome to informativeness, which is in line with the observed increase in loss learning rate adjustment. The effect in weight adjustment correlated with the effect in learning rate adjustment across participants, indicating that these two measures might capture similar behaviour. The effect of low mood on learning rate adjustment was not clearly supported by the regression results. However, decreased loss learning rate adjustment has previously been reported in the literature (16, 17). Replication studies are needed to assess the reliability of the findings observed in this study.

## 5 Conclusions

Our findings indicate that bifrontal tDCS might normalise the adjustment of learning rates to informativeness in individuals with depressive symptoms. This study therefore provides preliminary evidence that tDCS might normalise aberrant reinforcement learning processes which are hypothesised to lead to negative biases in depression. This effect was only present if tDCS was applied *during*, but not if applied *before* task performance, indicating that the cognitive state during stimulation matters. This suggests that combining tDCS with a concurrent cognitive manipulation might increase the functional impact on cognitive processes and potentially on mood. In future studies, we aim to investigate whether improvements in learning rate adjustment transfer to other tasks and might ultimately lead to improvements in mood.

## Data Availability

All data produced in the present study are available upon reasonable request to the authors.

## Acknowledgements

VS was funded by a studentship from the Medical Research Council (MR/N013468/1). MJO was funded by a scholarship from the Medical Research Council. MB is supported by a Clinician Scientist Fellowship from the MRC (MR/N008103/1). JO’S is a Sir Henry Dale Fellow funded by the Royal Society and the Wellcome Trust (215451/Z/19/Z). The Wellcome Centre for Integrative Neuroimaging is supported by core funding from the Wellcome Trust (203139/Z/16/Z and 203139/A/16/Z). This project was supported by the NIHR Oxford Health Biomedical Research Centre (NIHR203316). The views expressed are those of the author(s) and not necessarily those of the NIHR or the Department of Health and Social Care. For the purposes of open access the authors have applied a CC BY public copyright license to any Author Accepted Manuscript version arising from this submission.

The authors would like to acknowledge the use of the University of Oxford Advanced Research Computing (ARC) facility in carrying out this work. http://dx.doi.org/10.5281/zenodo.22558

