## Supplementary material for "Bifrontal transcranial direct current stimulation normalises learning rate adjustment in low mood"

### 1 Model comparison

#### 1.1 Alternative models

**Model 1.** This model used a modified version of a Rescorla-Wagner updating rule in which the probability of an outcome being associated with shape A was modelled separately for win and loss outcomes. The separate probabilities of a win or loss being associated with shape A, *rwin_(i+1)_* and *rloss_(i+1)_,* were modelled with separate learning rates for wins and losses (*αwin* and *αloss*):

$${rwin}_{\left( i+1 \right)}= {rwin}_{(i)}+ \alpha win *({winout}_{(i)}- {rwin}_{(i)})$$

$${rloss}_{\left( i+1 \right)}= {rloss}_{(i)}+ \alpha loss *({lossout}_{(i)}- {rloss}_{(i)})$$

A Softmax function was used to transform the trial-wise probability estimates into choice probabilities:

$${P(choice=A)}_{(i)}= \frac{1}{1+ {exp}^{(-\beta* \left( {rwin}_{\left( i \right)}- {rloss}_{\left( i \right)}+t \right))}}$$

In contrast to Model 6, this model only included one inverse temperature parameter ($\beta$), and a tendency parameter *t* to account for a potential tendency of choosing one shape over the other.

**Model 2.** Alternatively, participants might over- or underestimate the probability of wins being associated with shape A vs. losses being associated with shape A. Therefore, the Softmax function in model 2 contained two separate inverse temperatures for wins and losses that independently scaled the estimated probability of wins and losses.

$$P_{(choice=A\left( i \right))}= \frac{1}{1+\exp^{-({\beta win*rwin}_{\left( i \right)}-{\beta loss*rloss}_{\left( i \right)})}}$$

The probability estimates ${rwin}_{\left( i+1 \right)}$ and ${rloss}_{\left( i+1 \right)}$ were calculated as in model 1.

**Model 3.** While models 1 and 2 assume that participants maintain separate probability estimates for wins and losses, participants might simply learn the overall value of the shapes. In model 3, the overall value of shape A on trial *i+1,*${v^{A}}_{(i+1)}$, is calculated by

${v^{A}}_{(i+1)}=v_{(i)}^{A}+ a*({out}_{\left( i \right)}- v_{(i)}^{A}$)

In this model, the overall value of shape A on trial *i* is updated by the product of a single learning rate parameter $a$ and the prediction error $({out}_{\left( i \right)}- v_{(i)}^{A}$). ${out}_{\left( i \right)}$ represents the overall outcome and is coded as 1, -1 or 0, depending on whether shape A was associated with a win, a loss, both or neither outcome. The value estimates of the two shapes were transformed into action probabilities using a Softmax function with one inverse temperature parameter.

**Model 4.** Model 4 is identical to model 1 but does not include a tendency parameter in the Softmax function:

$${P(choice=A)}_{(i)}= \frac{1}{1+ {exp}^{(-\beta* \left( {rwin}_{\left( i \right)}- {rloss}_{\left( i \right)} \right))}}$$

**Model 5.** In model 5, the estimated probabilities of the win or loss being associated with shape A are updated using the same learning rate parameter $\alpha:$

$${rwin}_{\left( i+1 \right)}= {rwin}_{(i)}+ \alpha*({winout}_{(i)}- {rwin}_{(i)})$$

$${rloss}_{\left( i+1 \right)}= {rloss}_{(i)}+ \alpha*({lossout}_{(i)}- {rloss}_{(i)})$$

The estimated probabilities are transformed into choice probabilities using the same Softmax function as in model 2, i.e. separate inverse temperature parameters for win and loss outcomes.

**Model 6.** Model 6 is similar to model 2, but includes separate inverse temperature parameters for win and loss outcome which are centred around zero:

$P_{(choice=A\left( i \right))}= \frac{1}{1+\exp^{(-\left( {\beta win*(rwin}_{\left( i \right)}- 0.5 \right)-\left( {\beta loss*(rloss}_{\left( i \right)}- 0.5 \right))}}$

This model accounts for differential sensitivity to win or loss outcomes.

#### 1.2 Parameter estimation in STAN

Parameters for the constant models were estimated in STAN (1) which uses a Markov Chain Monte Carlo (2) algorithm to sample from the Bayesian posterior distribution. 4 chains were run for 10,000 iterations (adapt_delta = 0.99, max_treedepth = 12). The prior mean was set to 0 for the inverse temperature (log-transformed), and to -0.5 for learning rates (quantile-transformed) which is the average value observed in the block-wise model. The standard deviation for priors was set to 1. The Gelman-Rubin statistics (R-hat) was below 1.1 for all estimates, indicating good convergence (3).

#### 1.3 Model comparison block-wise models

Model comparison for the sample of participants with low mood was performed using the BIC summed up over the six task blocks, and averaged across sessions and participants. The parameters of each model are summarised in Table S1. In our previous study in healthy volunteers (4), model 1 fitted the data best. In line with our previous study, model 1 fitted the data best (Figure S2). Parameters from model 1 were therefore used in all further analyses using the block-wise model.

Table S1. Number of parameters *per block* for the block-wise models.

| Model | Learning rate parameters | Inverse temperature parameters | Notes |
| --- | --- | --- | --- |
| 1 | 2 | 1 | Tendency parameter |
| 2 | 2 | 2 | Inverse temperature parameters scale estimated outcome probabilities |
| 3 | 1 | 1 | Overall value |
| 4 | 2 | 1 | Identical to model 1 without tendency parameter |
| 5 | 1 | 2 | Estimated win and loss probabilities are updated using the same learning rate |
| 6 | 2 | 2 | Inverse temperature parameter scale centered estimated outcome probabilities |

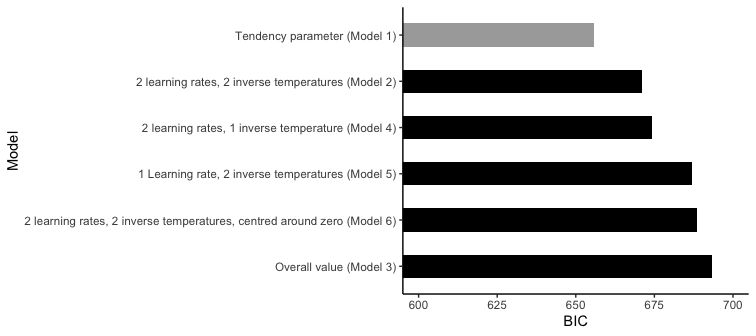

*Figure S2.* Model comparison of the block-wise models for the sample of participants with low mood. As in the previous study, model 1 fitted the data best.

To test whether our procedure of parameter estimation can reliably reproduce parameter values from the winning model, parameter recovery was performed for model 1. First, choices were simulated from model 1 using 1000 combinations of parameter values. For each of the four parameters, 10 equally spaced values covering the entire parameter range considered in the estimation procedure were used for simulating choices. To test up to which level increases in inverse temperature still produce differential choice behaviour, we calculated how many choices differed with each increase in inverse temperature. We found that the influence of increases in inverse temperature declined very steeply, leading to less than one different choice on average for the highest inverse temperature values. For parameter recovery, the range of the inverse temperature was therefore restricted to a maximum of 44.5 (~2 choices were different from previous value). 10 equally spaced parameter values up to 44.5 were then used for parameter recovery. Model 1 was than fitted to the simulated choices and the resulting parameter estimates were compared to the actual parameter estimates used for simulating the choices. Overall, the results indicate that the estimated parameters were reasonably close to the actual parameter values used for simulation. More details are included in Figure S3.

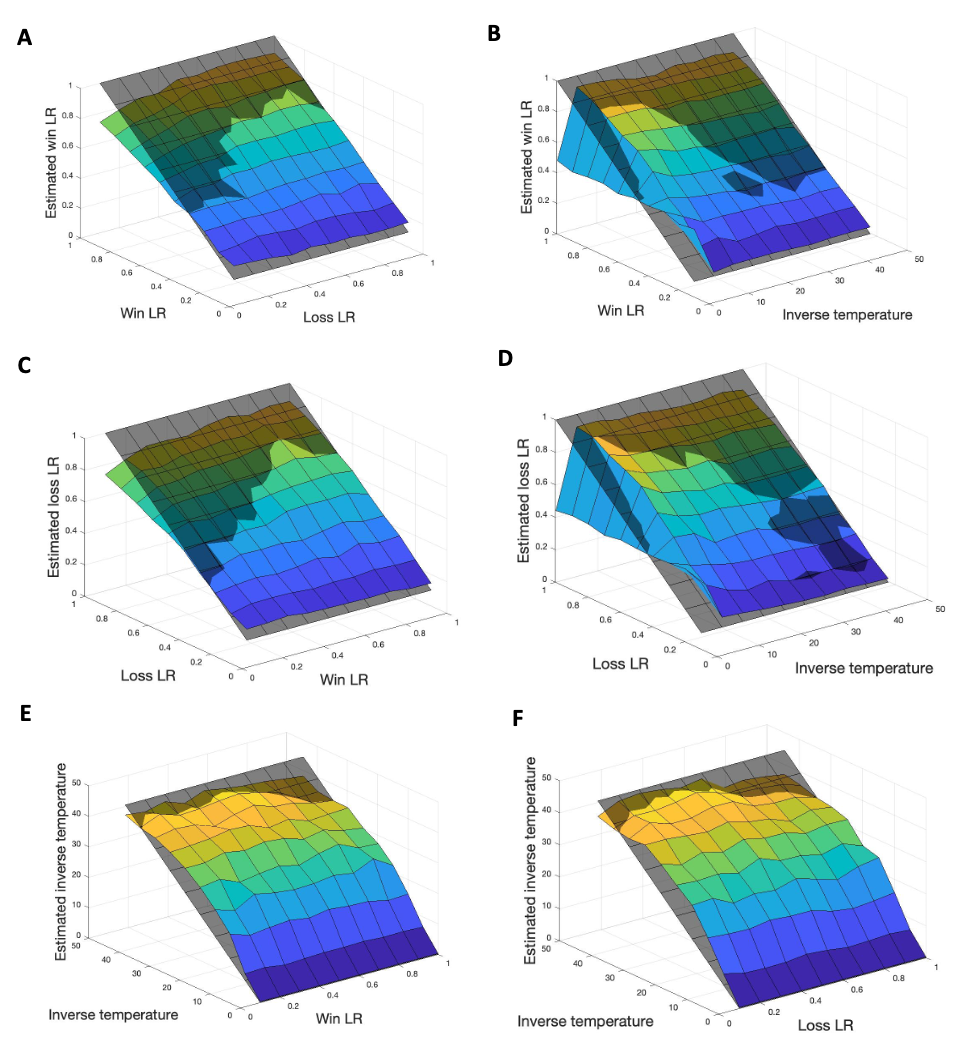

*Figure S3.* Parameter recovery for model 1. Recovery of win learning rate depending on loss learning rate (A) or inverse temperature (B). Recovery of the loss learning rate depending on the win learning rate (C) or inverse temperature (D). Recovery of the inverse temperature depending on the win learning rate (E) or loss learning rate (F). The learning rate for one outcome tends to be underestimated if the learning rate for the other outcome is low (A and C). Learning rates cannot reliably be estimated if the inverse temperature is very low (B and D).

#### 1.4 Model comparison constant models

Model comparison for the constant models was performed across all participants from our previous study in healthy participants and the sample of participants with low mood. The parameters of each model are summarised in Table S2. The BIC was averaged across sessions and participants. Model 6 provided the best fit (Figure S4). The parameter estimates derived from Model 6 were therefore used for all analyses of the constant model.

Table S2. Number of parameters *per session* for the constant models.

| Model | Learning rate parameters | Inverse temperature parameters | Notes |
| --- | --- | --- | --- |
| 1 | 12 | 1 | Tendency parameter (one per block) |
| 2 | 12 | 2 | Inverse temperature parameters scale estimated outcome probabilities |
| 3 | 6 | 1 | Overall value |
| 4 | 12 | 1 | Identical to model 1 without tendency parameter |
| 5 | 6 | 2 | Estimated win and loss probabilities are updated using the same learning rate |
| 6 | 12 | 2 | Inverse temperature parameter scale centered estimated outcome probabilities |

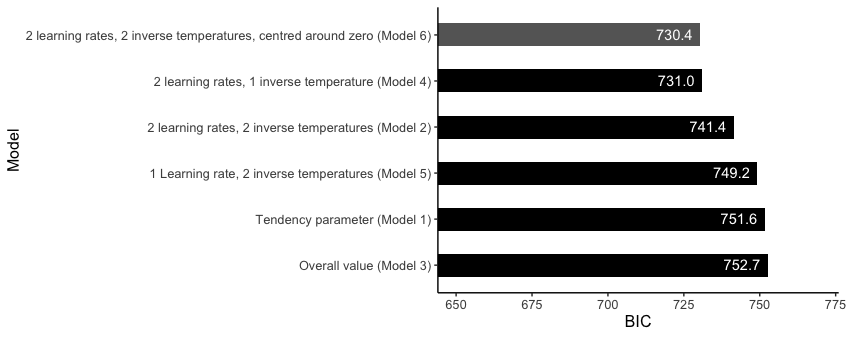

*Figure S4.* Model comparison. According to the BIC, Model 6 fitted the data best.

Parameter recovery was performed based on the parameter estimates obtained from model 6 for all participants from the previous study in healthy individuals and participants with low mood. Parameter estimates for each participant and session were used to simulate choices in the task. Model 6 was then fitted to the simulated datasets, and the obtained parameter estimates were compared to the original parameters. Estimated and actual parameters were highly correlated, with correlation coefficients between .73 and .86 (Figure S5).

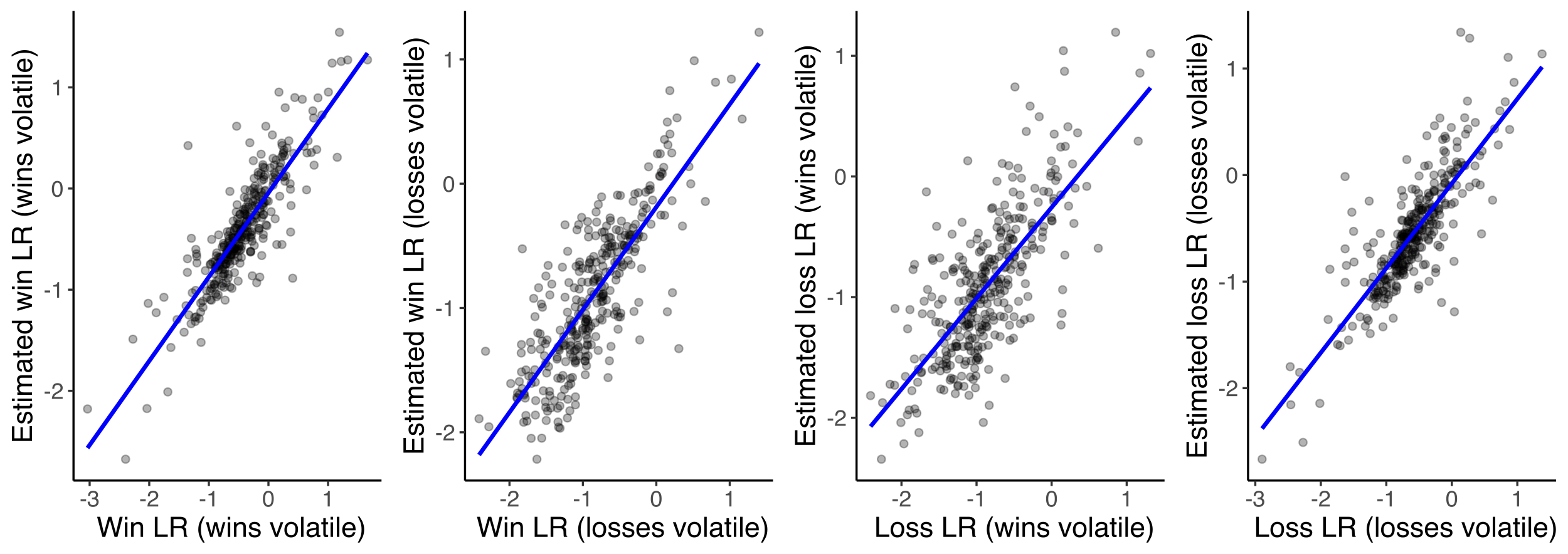

r = .86

r = .73

r = .81

r = .82

*Figure S5.* Parameter recovery for model 6 for the parameters of interest, i.e. win and loss learning rates in the wins-volatile and losses-volatile blocks. All parameter estimates were highly correlated with the original parameter values. Parameter recovery was better for learning rates of the volatile outcome (win learning rate in the wins-volatile block, and loss learning rate in the losses-volatile block).

#### 1.5 Trial-by-trial model fit

Trial-by-trial model prediction for the best block-wise model and constant model averaged across participants from both samples is shown in Figure S6.

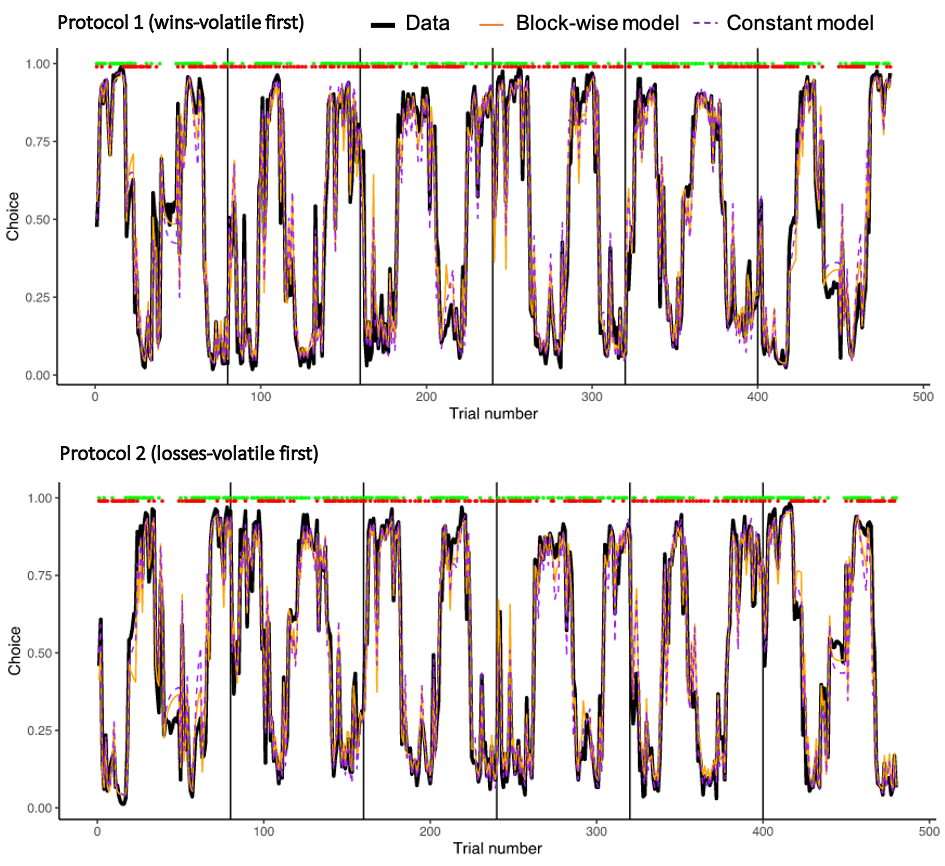

*Figure S6*. Comparison of the trial-wise model prediction of the block-wise model (model 1, orange line) and constant model (model 6, dashed purple line) with the actual data (thick black line). The thick black line indicates for each trial the percentage of participants who chose shape A. The other two lines represent the trial-by-trial likelihood of choosing shape A, derived from the parameter estimates from all participants. The trial-wise likelihood was averaged across all participants and sessions. Green and red points indicate for each trial whether the win was associated with shape A (green dot) or shape B (no green dot), and whether the loss was associated with shape A (red dot) or shape B (no red dot).

### 2 Supplementary methods

#### 2.1 Non-computational outcome measures

Non-computational measures included total winnings, number of wins received, number of losses received, win-stay probability (probability of choosing the same option on the next trial after receiving a win outcome), no-loss-stay probability (probability of choosing the same option on the next trial after not receiving a loss outcome), loss-switch probability (probability of choosing the other option on the next trial after receiving a loss outcome) and no-win-switch probability (probability of choosing the other option on the next trial after not receiving a win outcome).

As a non-computational measure of positive bias, we included the proportion of win-driven choices. This measure was based on choices that participants made after a trial in which they received both a win and a loss outcome in response to their chosen shape. These trials are of particular interest because they can differentiate between two opposite strategies: if participants are primarily trying to maximise the number of wins they receive, then on the next trial they should choose the same shape, because it was associated with the win (both outcomes) on the previous trial. However, if participants are mainly trying to avoid losses, on the next trial they should instead pick the alternative shape that was not associated with the loss (neither outcome) on the previous trial. The proportion of “win-driven choices” is therefore the number of such trials in which participants choose the shape that was associated with both outcomes, divided by the total number of trials in which the win and loss were associated with the same shape.

All dependent variables (apart from stay probabilities) were analysed in mixed ANOVAs, with the main factor of interest being the factor Sample (general population vs. low mood). All analyses included the between-subject factor Block Order (wins-volatile first vs. losses-volatile first) and the within-subject factors Volatility (both-volatile, wins-volatile and losses-volatile) and Time (first half vs. second half). Win-stay probability and no-loss-stay probability were analysed with Wilcoxon rank-sum tests (effect of low mood) or Wilcoxon signed-rank tests (effect of tDCS) due to violations of the normality assumption.

#### 2.2 Outlier removal

All analyses were repeated after removing outliers. A datapoint was identified as outlier if it was more than 1.5 times the interquartile range below the first or above the third quartile. For each outcome measure, outliers were removed separately for the levels of the factors of interest (i.e. separately for the general population and low mood sample, and win and loss outcomes where appropriate). For the analysis of the effect of real vs. sham tDCS, a datapoint was identified as outlier if the difference between real minus sham tDCS was more than 1.5 times the interquartile range below the first or above the third quartile. Statistics are reported for the entire dataset unless outlier removal had an impact on the results (for completeness, figures including outliers are included for all analyses that outlier removal had an impact on).

### 3 Detailed results: Low mood is associated with alterations in learning rate adjustment

Summary statistics for all computational parameters are provided in Table S4 – S15 (section 6).

#### 3.1 The effect of low mood on non-computational measures

Overall, participants suffering from low mood won less money in the task (Sample: *F*(1,79) = 8.3, *p* = .004), received fewer wins (Sample: *F*(1,79) = 8.0, *p* = 005), and more losses (Sample: *F*(1,79) = 4.5, *p* = .036)(Figure S7).

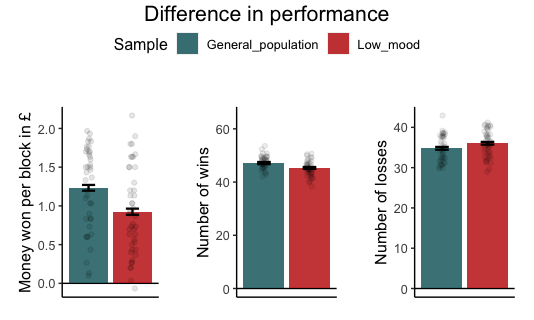

C

A

B

*Figure S7.* Effect of low mood on task performance. Participants in the low mood sample won less money (A) and received less wins (B) and more losses (C) compared to the general population sample.

Participants in the low mood sample switched more often (Sample: *F*(1,79) = 5.0, *p* = .02), were less likely to stay after receiving a win (win-stay probability: *W = 1118, p* = .018) or after not receiving a loss (no-loss-stay probability: *W* = 1136, *p* = .012). There was no effect of Sample on the no-win-switch probability (Sample: *F*(1,79) = 2.7, *p* = .10) but a trend towards higher loss-switch probability in the low mood sample (Sample: *F*(1,78) = 3.2, *p* = .076)(Figure S8).

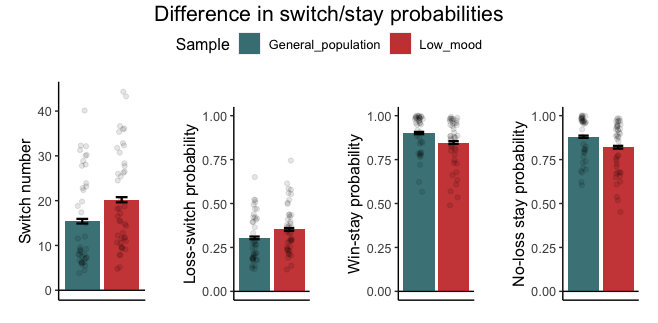

A

B

C

D

Figure S8. Effect of low mood on switch behaviour. Participants in the low mood sample switched more often (A), showed a trend towards higher loss-switch probability (B), had lower win-stay probabilities (C), and lower no-loss-stay probabilities (D).

Participants with low mood did not differ from the general population sample in the proportion of win-driven choices (Sample: *F*(1,79) = 0.02, *p* = .87). Across groups, BDI scores were negatively correlated with total winnings (*r* = -.23, *t*(79) = 2.0, *p* = .042) and the number of received wins (*r* = ‑.35, *t*(77) = -3.3, *p* = .001). However, within the two samples no significant relationship between BDI or trait anxiety scores with any of the non-computational measures was observed (all *p* > .26).

#### 3.2 The effect of low mood in the block-wise model

To test our first hypothesis that individuals with low mood would show increased punishment vs. reward learning, learning rate parameters derived from model 1 were compared between the two samples. The low mood sample showed comparable learning rates to the general population sample (Sample: *F*(1,79) = 1.9, *p* = .16; Sample x Valence: *F*(1,79) = 0.1, *p* = .66); Sample x Valence x Volatility: *F*(2,158) = 0.1, *p* = .86)(Figure S9A).

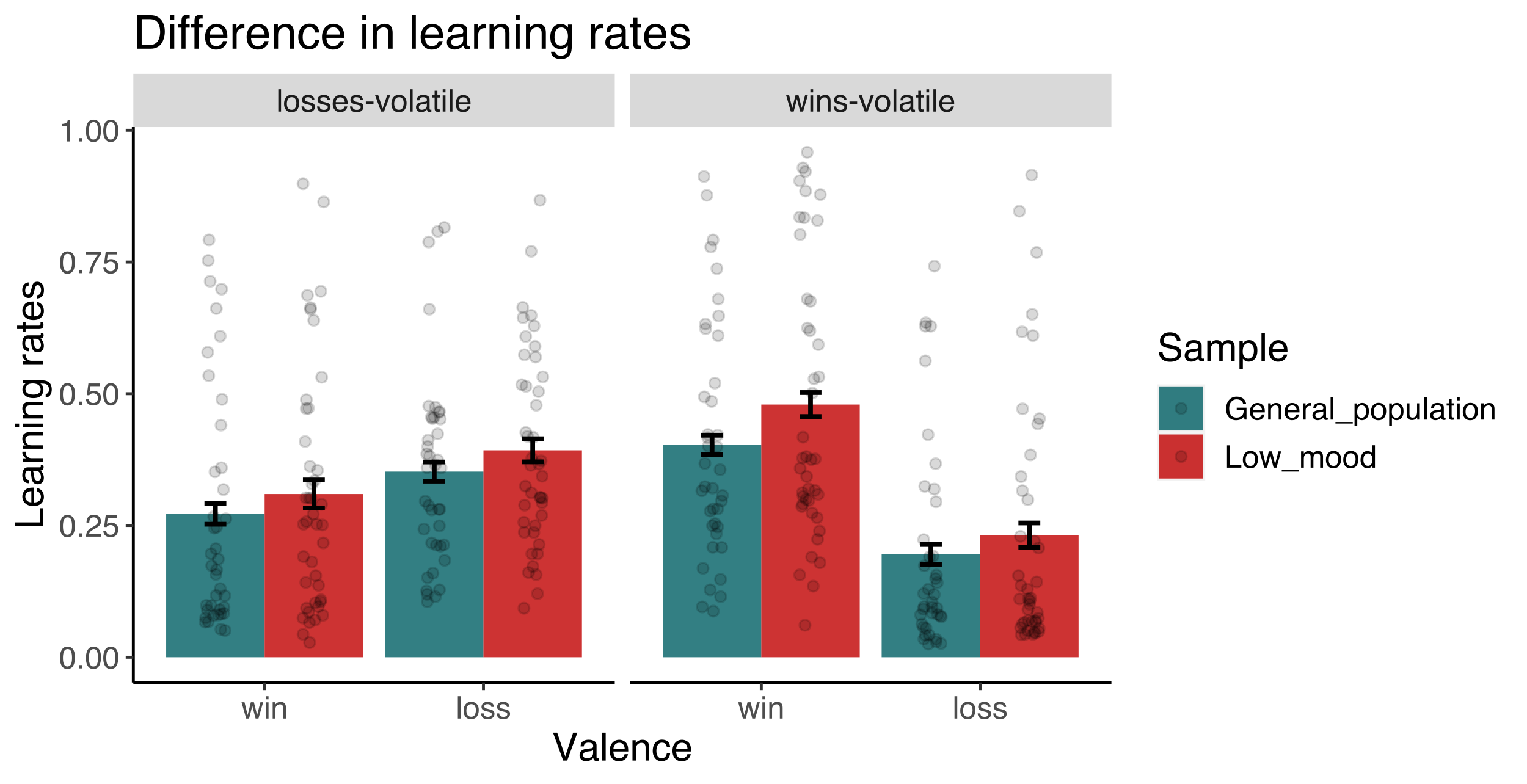

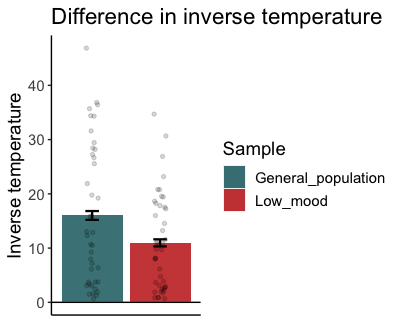

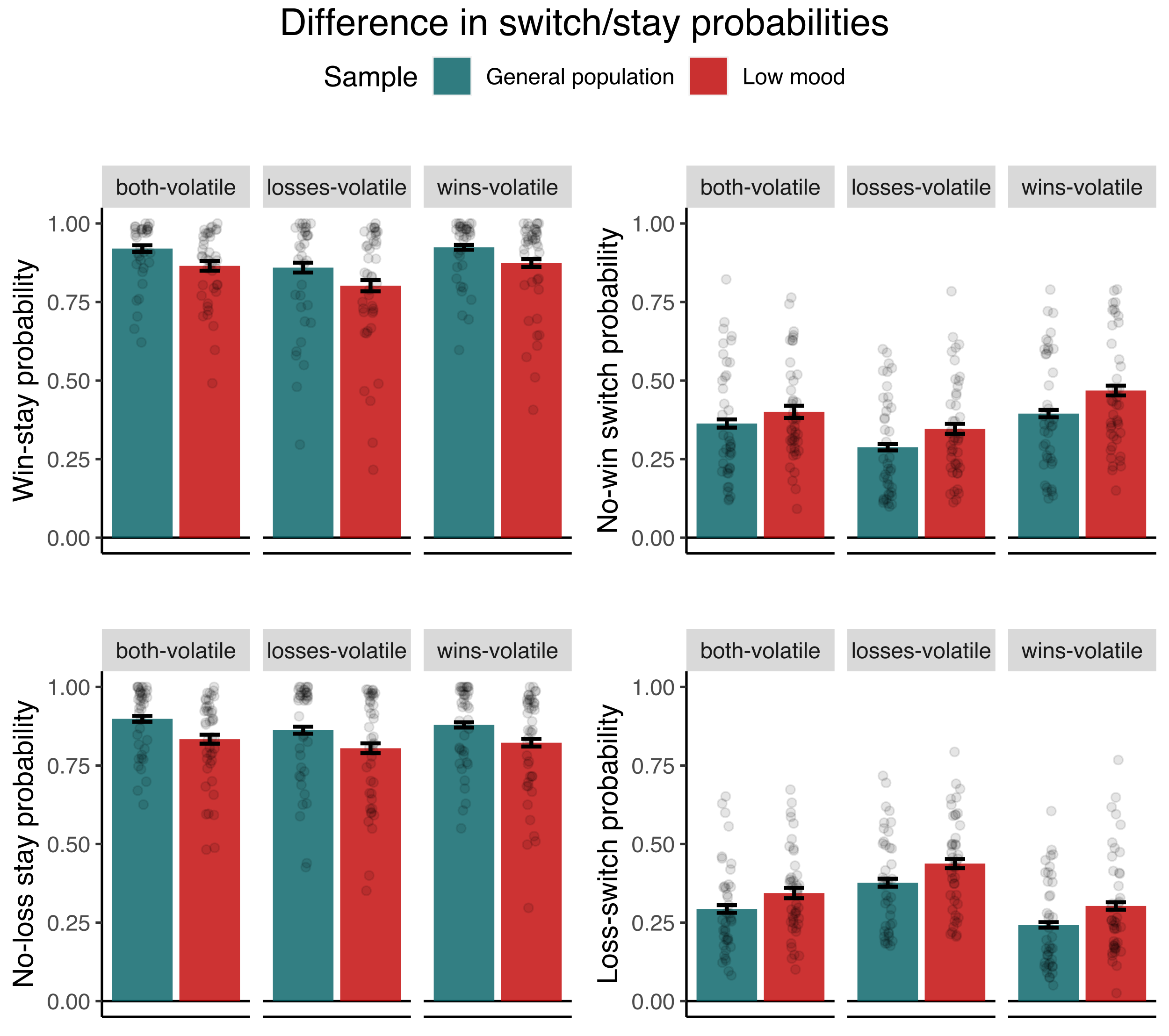

B

A

Figure S9. Effect of low mood on learning rates and inverse temperature in the block-wise model. The low mood sample had comparable win and loss learning rates (A), but lower inverse temperature estimates (B) compared to the general population sample.

To test our second hypothesis that individuals with would low mood would adjust their learning rates in response to volatility to a smaller extent than healthy individuals, an ANOVA on *learning rate adjustment* was conducted. There was no main effect of Sample (*F*(1,79) = 0.1, *p* = .65) or interaction effect between Sample and Valence (*F*(1,79) = 0.9, *p* = .34). *Learning rate adjustment bias* (i.e. *loss learning rate adjustment* minus *win learning rate adjustment*) did not differ either between the two samples (*F*(1,79) = 0.9, *p* = .34)(Figure S10).

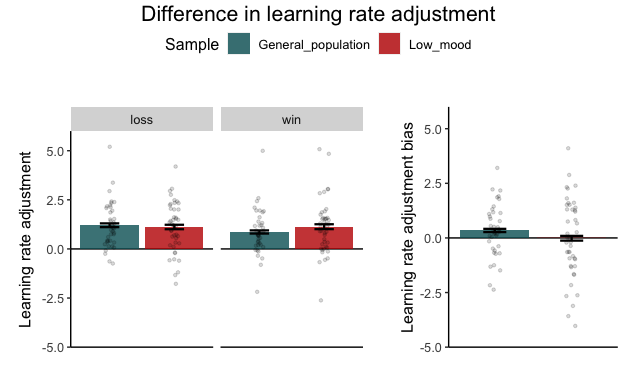

A

B

Figure S10. Effect of low mood on learning rate adjustment. Participants suffering from low mood did not differ from the general population sample in learning rate adjustment (A) or learning rate adjustment bias (B).

To test whether individuals with low mood differed from healthy individuals in choice randomness, the inverse temperature estimates were compared between the two samples. Individuals with low mood had lower inverse temperature estimates, i.e. were more random in their choice behaviour (Sample: F(1,79) = 4.9, p = .029)(Figure S9B).

No computational measure was significantly correlated with the BDI or trait anxiety score (all *p* > .11).

#### 3.3 The effect of low mood in the constant model

To test our first hypothesis that individuals with low mood would show increased punishment vs. reward learning, learning rate estimates were analysed in an ANOVA with Sample and Valence as the factors of interest. Contrary to this hypothesis, there was no main effect of Sample (Sample: *F*(1,79) = 0.5, *p* = .46). There was a trend towards an interaction between Sample and Valence (Sample x Valence: *F*(1,79) = 3.7, *p* = .056), but the effect of Sample was neither significant for win nor for loss learning rates (win learning rate: *F*(1,79) = 0.06, *p* = .79); loss learning rate: *F*(1,79) = 2.4, *p* = .12). Win and loss learning rates were not significantly correlated with the BDI or trait anxiety score (all *p* > .29).

To test our second hypothesis that individuals with low mood would adjust their learning rates to a smaller extent than healthy individuals, learning rate adjustment was analysed in an ANOVA with Sample and Valence as main factors of interest. There was a Sample x Valence interaction (after removal of 7 outliers: *F*(1,72) = 4.4, *p* = .038; before outlier removal: *F*(1,79) = 4.0, *p* = .046, see Figure S11). Post-hoc tests indicated that there was no significant main effect of Sample on win learning rate adjustment (*F*(1,72) = 1.3, *p* = .24) but a trend towards lower loss learning rate adjustment in the sample with low mood (*F*(1,72) = 3.2, *p* = .076)(Figure 4B, main text). However, in line with the interaction effect of Sample and Valence on learning rate adjustment, there was a main effect of Sample on the combined measure of *learning rate adjustment bias* (after outlier removal: *F*(1,72)= 4.4, *p* = .038, *Cohen’s* *d* = 0.44; before outlier removal: *F*(1,79) = 4.0, *p* = .046)(Figure 4C, main text). Individuals with low mood showed a lower *learning rate adjustment bias*. That is, while the general population adjusted their win and loss learning rates to a similar extent (one-sample t-test against zero of the learning rate adjustment bias (general population): *t*(35) = -1.3, *p* = 0.17), individuals with low mood adjusted their loss learning rate significantly less than their win learning rate (one-sample t-test against zero of the learning rate adjustment bias: *t*(39) = -3.2, *p* = .002).

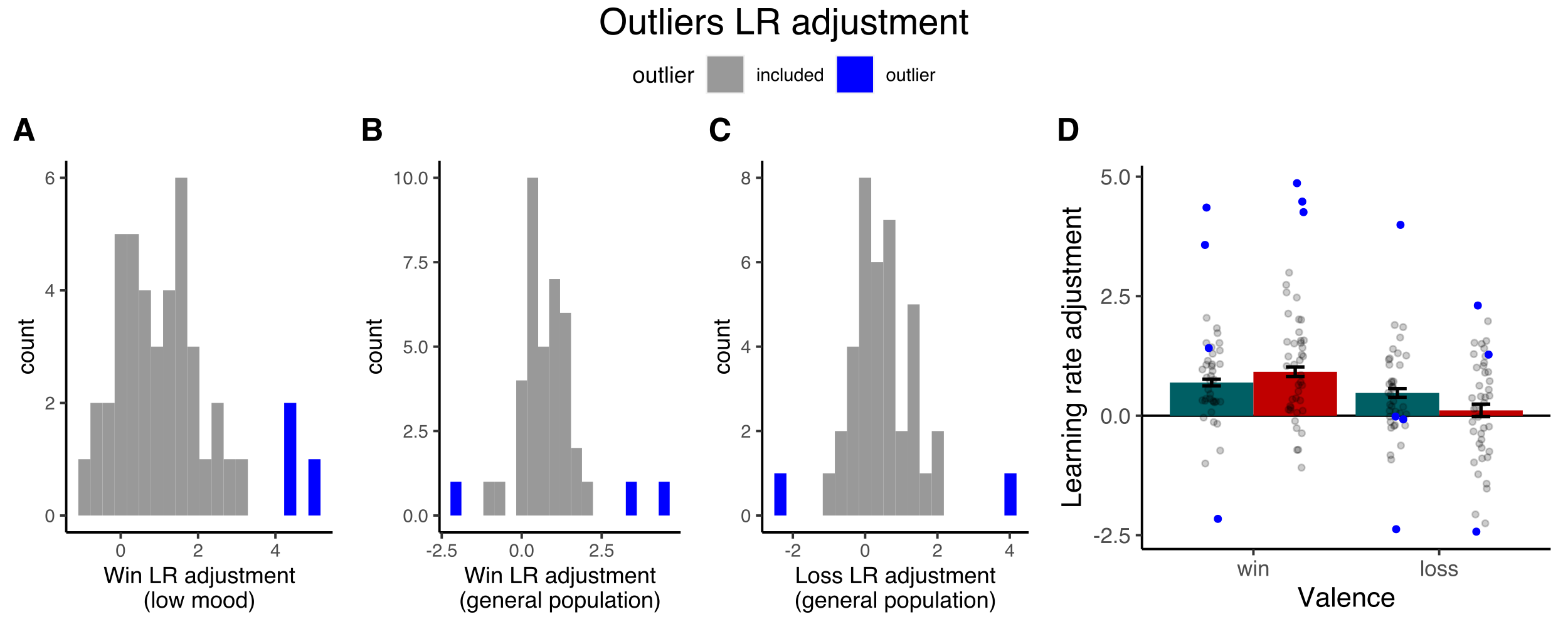

*Figure S11.* Outlier removal for *learning rate adjustment.* Outliers were identified separately for win and loss learning rate adjustment, and for the low mood and general populations sample (low mood: 3 outliers, general population: 4 outliers). (A) Outliers for win learning rate adjustment in the low mood sample, (B) outliers for win learning rate adjustment in the general population sample, (C) outliers for loss learning rate adjustment in the general population sample (there were no outliers for loss learning rate adjustment in the low mood sample). (D) shows the difference in learning rate adjustment between the samples including outliers (see Figure 4 in the main text). Outliers are shown in blue. Due to the within-subject design, subjects who were classified as outliers in (A) or (B) were removed entirely from this analysis.

To test for a linear relationship between mood and learning rate adjustment, BDI and trait anxiety scores were examined for correlation with learning rate adjustment. Across groups, BDI scores correlated negatively with the learning rate adjustment bias, i.e. individuals with more severe depressive symptoms showed a more negative *learning rate adjustment bias* (r = -.25, *t*(75) = -2.2, *p* = .027). This is in line with the group differences observed in the ANOVA. In line with this, there was a non-significant negative correlation between BDI scores and loss learning rate adjustment (r = -.18, *t*(75) = -1.5, *p* = .11) and a trend towards a positive correlation between BDI scores and win learning rate adjustment (r = .20, *t*(73) = 1.8, *p* = .070). However, the same correlations conducted within each group were not significant (see Figure S12), indicating that the correlation between BDI and learning rate adjustment was driven by the group difference. On a descriptive level, the correlations in the low mood sample (which has a higher variance in the BDI scores than the general population sample) were in the same direction as the correlations across the two samples.

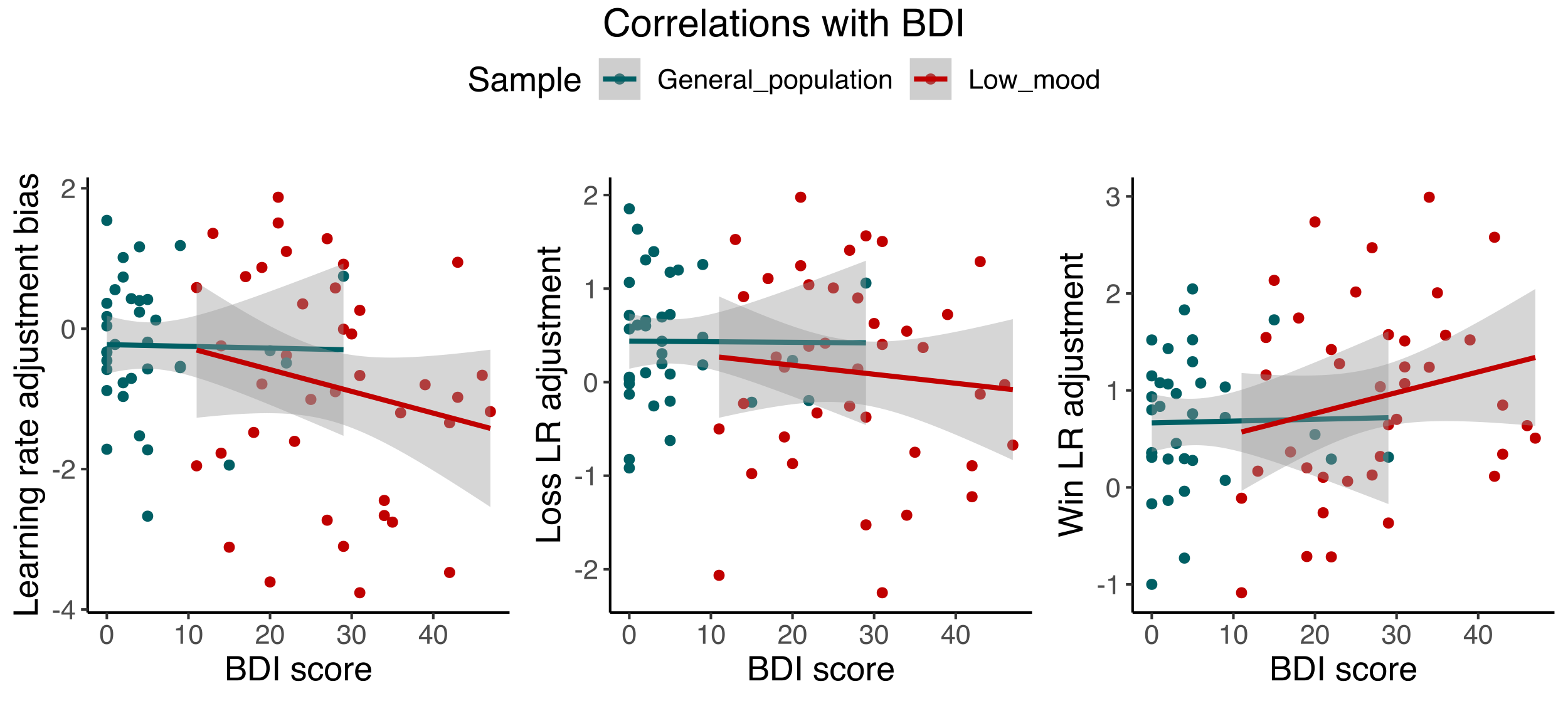

A

B

C

r = -.19, *p* = .22

r = -.01, *p* = .91

r = -.09, *p* = .57

r = -.006, *p* = .97

r = .21, *p* = .18

r = .01, *p* = .91

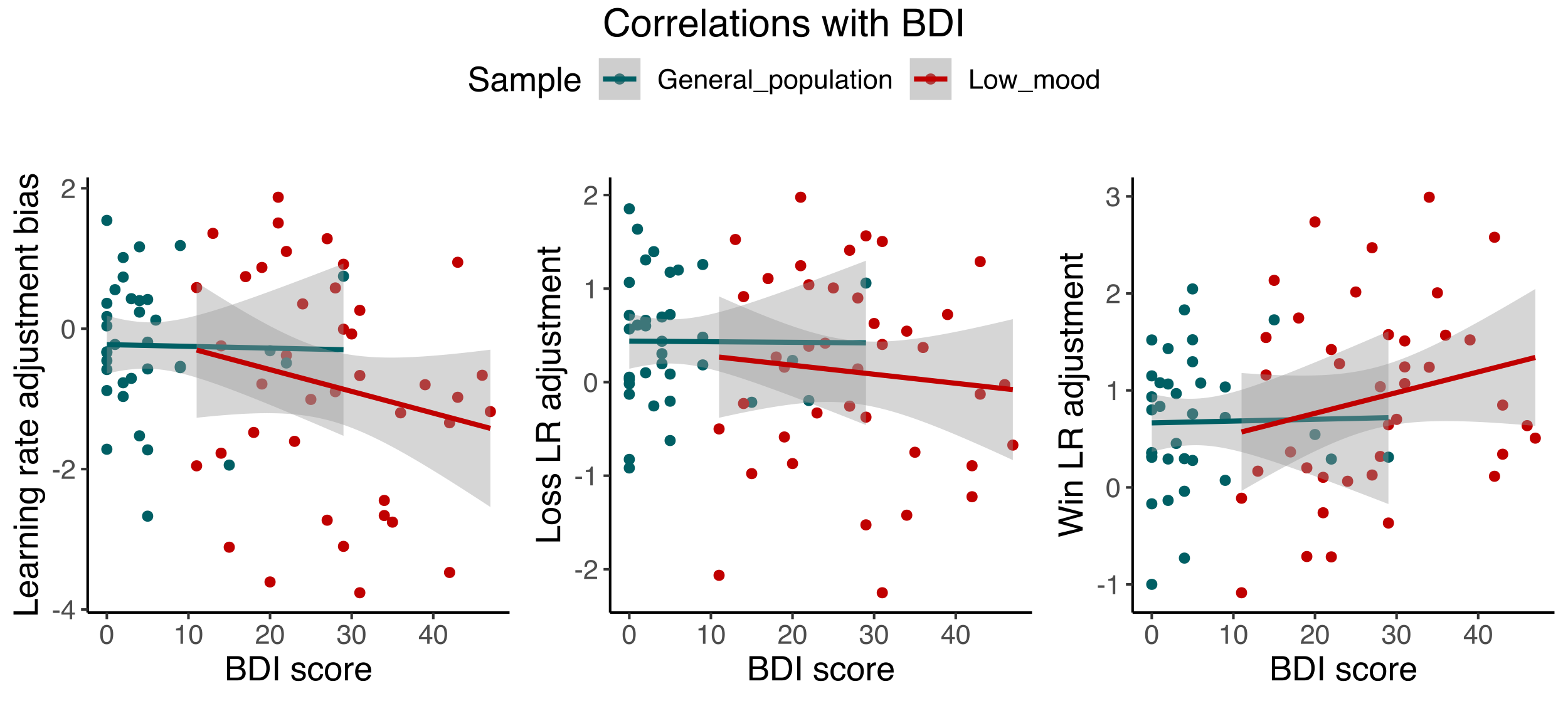

Figure S12. Correlations between BDI score and learning rate adjustment bias (A), loss learning rate adjustment (B) and win learning rate adjustment (C).

To test whether individuals with low mood differed from healthy individuals in choice randomness, the inverse temperature estimates for wins and losses were analysed in an ANOVA with Sample and Valence as factors of interest. There was a marginally significant main effect of Sample on the inverse temperature estimates (*F*(1,78) = 3.9, *p* = .050). Participants with low mood showed lower inverse temperatures for wins and losses, independent of their valence (Sample x Valence: *F*(1,78) = 0.17, *p* = .67). We tested for correlations of the BDI or trait anxiety score with the inverse temperature estimates for wins, losses or averaged across both outcomes. Across groups, there was a significant negative correlation between the BDI score and win inverse temperature (*r* = -.21, *t*(80) = -1.9, *p* = .047) which became non-significant after removing the most extreme datapoint (*r* = -.14, *t*(79) = -1.3, *p* = .19). There was no significant correlation between BDI or trait anxiety score with the loss inverse temperature or average inverse temperature, neither across nor within groups (all *p* > .13).

### 4 Detailed results: Online bifrontal tDCS normalises learning rate adjustment

#### 4.1 The effect of tDCS on non-computational measures

To test whether bifrontal tDCS applied during task performance had an effect on non-computational measures, we analysed total winnings, stay and switch probabilities and proportion of win-driven choices during stimulation (block 2 and 3) in separate ANOVAs. There was no effect of tDCS on any non-computational measure (Money won: tDCS: *F*(1,40) = 0.2, *p* = .60, tDCS x Volatility: *F*(1,40) = 0.5, *p* = .48); win-stay probability: tDCS: *V* = 363, *p* = .71; no-loss-stay: tDCS: *V* = 369, *p* = .43; no-loss-switch probability: tDCS: *F*(1,40) = 0.3, *p* = .55, tDCS x Volatility: *F*(1,40) = 2.5, *p* = .12; no-win-switch: tDCS: *F*(1,40) = 0.02, *p* = .87; proportion of win-driven choices: tDCS: *F*(1,40) = 0.6, *p* = .43, tDCS x Volatility: *F*(1,40) = 1.1, *p* = .28).

We further tested whether bifrontal tDCS applied *before* task performance had an effect on non-computational measures. TDCS applied before task performance had no effect on any non-computational measure (Money won: tDCS: *F*(1,43) = 0.4, *p* = .52, tDCS x Volatility: *F*(1,43) = 0.008, *p* = .92; win-stay probability: tDCS: *V* = 416, *p* = .85; no-loss-stay probability: tDCS: *V* = 381, *p* = .18; loss-switch probability: tDCS: *F*(1,43) = 0.2, *p* = .62, tDCS x Volatility: *F*(1,43) = 0.001, *p* = .96; no-win-switch probability: tDCS: *F*(1,43) = 0.23, *p* = .62, tDCS x Volatility: *F*(1,43) = 0.02, *p* = .88; proportion of win-driven choices: tDCS: *F*(1,43) = 1.5, *p* = .22, tDCS x Volatility: *F*(1,43) = 0.2, *p* = .65).

##### 4.2 The effect of tDCS in the constant model

To test our main hypothesis that tDCS would selectively increase the learning rate from wins, we ran an ANOVA with tDCS Condition and Valence as the main factors of interest. There was no significant main effect of tDCS nor an interaction between tDCS Condition and Valence, indicating that tDCS had no effect on learning rates (main effect of tDCS Condition: *F*(1,37) = 0.3, *p* = .86); tDCS Condition x Valence: *F*(1,37) = 0.26, *p* = .60)(Figure 5A, main text).

To test for a potential effect of tDCS on learning rate adjustment, we ran an ANOVA with learning rate adjustment as dependent variable. There was no main effect of tDCS Condition (*F*(1,34) = 0.1, *p* = .70), but there was a significant interaction effect between tDCS Condition and Valence (after removal of three outliers: *F*(1,34) = 7.47, *p* = .009; before outlier removal: (*F*(1,37) = 3.7, *p* = .059), see Figure S13). Real compared to sham tDCS led to an increase in loss learning rate adjustment (*F*(1,34) = 6.0, *p* = .018, *Cohen’s dz* = 0.46), and to a marginally significant decrease in win learning rate adjustment (*F*(1,34) = 4.0, *p* = .051, *Cohen’s dz* = ‑0.48) (Figure 5B, main text). In line with this, real vs. sham tDCS had a significant effect on the learning rate adjustment bias (*F*(1,34) = 7.4, *p* = .009, *Cohen’s dz* = 0.65; before outlier removal: *F*(1,37) = 3.7, *p* = .059). Real vs. sham tDCS increased the learning rate adjustment bias, i.e. shifted learning rate adjustment bias from wins towards losses (Figure 5C, main text). During sham tDCS, the learning rate adjustment bias was negative and significantly different from zero (*t*(37) = ‑2.1, *p* = .037), indicating that participants adjusted their loss learning rates significantly less than their win learning rate. During real tDCS, there was a trend towards a positive learning rate adjustment bias, i.e. towards higher loss than win learning rate adjustment (*t*(37) = 1.8, *p* = .067).

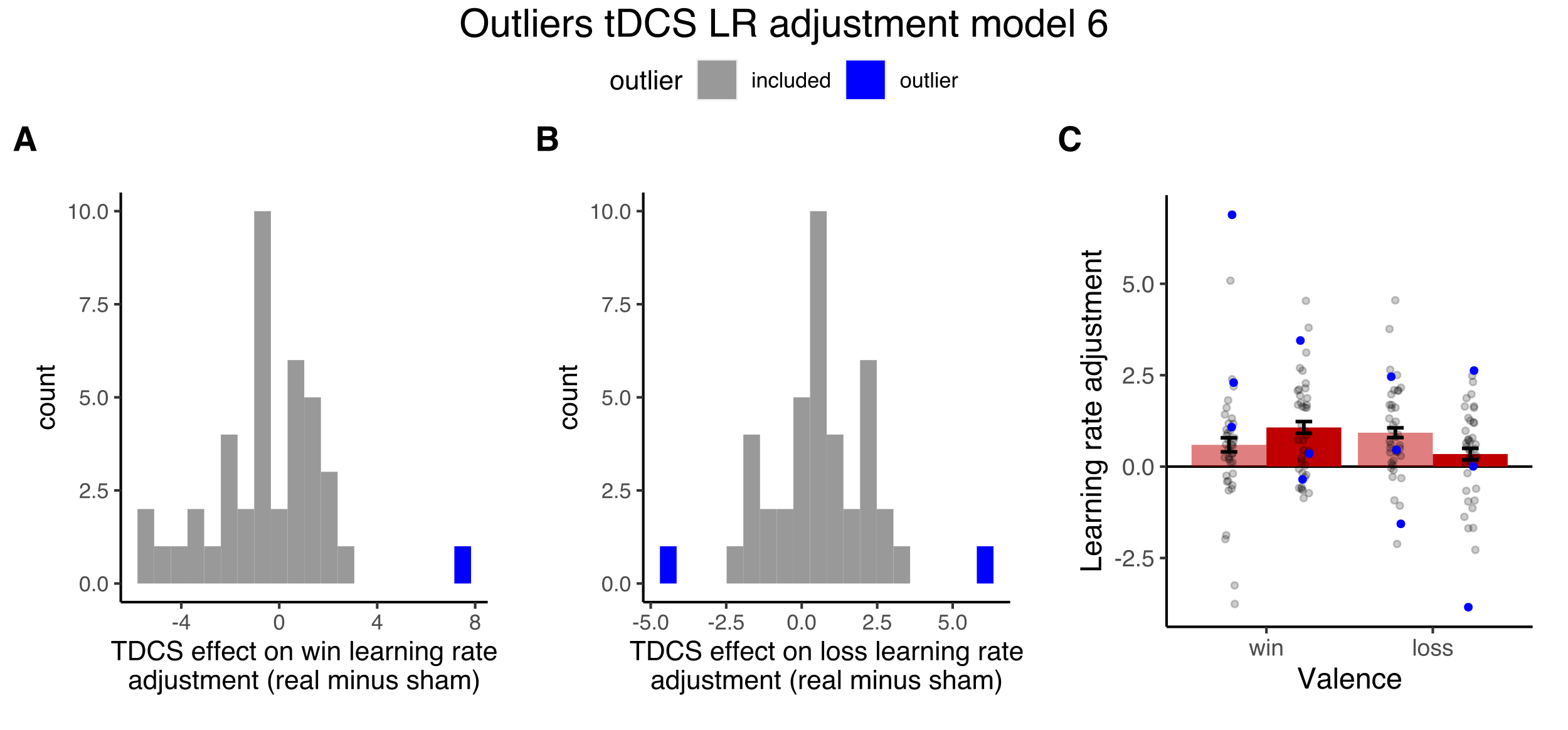

Figure S13. Outlier removal for learning rate adjustment based on the constant inverse temperature model (three outliers). Outliers were identified based on the difference in win (A) and loss learning rate adjustment (B) between real and sham tDCS. (C) shows the effect of tDCS on learning rate adjustment including outliers (see Figure 5B in the main text). Outliers are highlighted in blue. Due to the within-subject design, subjects who were classified as outliers in (A) or (B) were removed entirely from this analysis.

To test whether the effect of tDCS on learning rate adjustment outlasted the stimulation period, we ran the same ANOVAs with learning rate adjustment (or learning rate adjustment bias) as dependent variable in the task blocks after the stimulation period (block 3 and 4). There was no significant interaction between tDCS Condition and Valence (or main effect of tDCS Condition)(*F*(1,34) = 1.3, *p* = 0.24), indicating that the effect was confined to the period during stimulation (Figure S14).

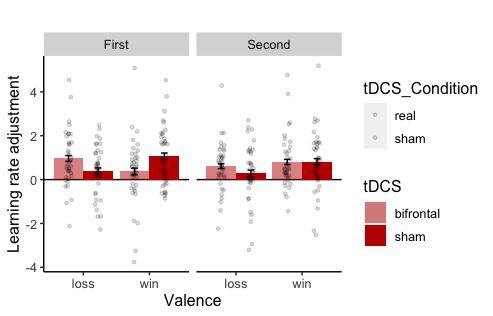

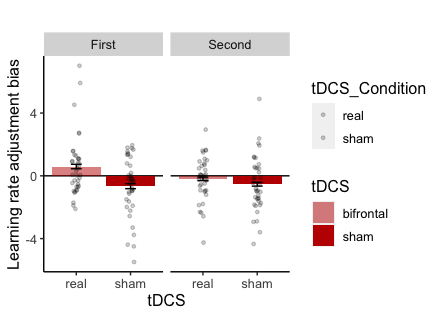

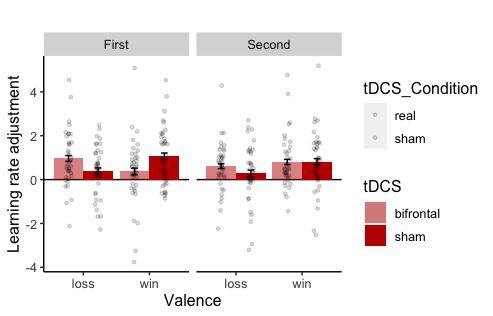

Figure 6.1

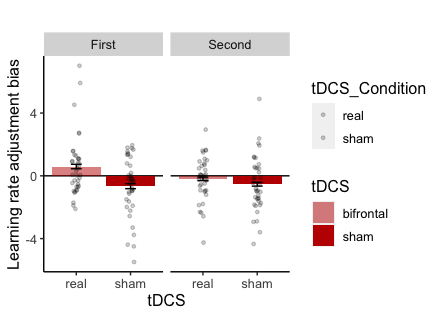

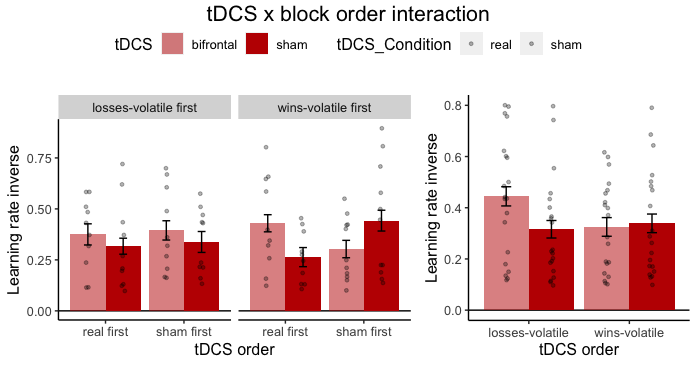

Figure S14. Effect of online bifrontal tDCS after the stimulation period. The effect of online bifrontal tDCS on learning rate adjustment (A) and learning rate adjustment bias (B) was only present during the stimulation time (First) but not after the stimulation had finished (Second).

As a control analysis, we also analysed the inverse temperature estimates which are unlikely to be affected by tDCS since they were estimated across the entire task. There was no effect of tDCS Condition on the inverse temperature (all *p* > .29).

We further tested our hypothesis that the effect of tDCS would be specific to the cognitive state during stimulation. In line with this hypothesis, offline tDCS had no significant effect on learning rates (tDCS: *F*(1,42) = 0.9, *p* = .32, tDCS x Valence: *F*(1,42) = 0.3, *p* = .55, tDCS x Volatility: *F*(1,42) = 0.001, *p* = .96, tDCS x Valence x Volatility: *F*(1,42) = 1,1, *p* = .30), *learning rate adjustment* (tDCS: *F*(1,42) = 1.1, *p* = .30, tDCS x Valence: *F*(1,42) = 0.001, *p* = .96), *learning rate adjustment bias* (tDCS: *F*(1,42) = 0.001, *p* = .96) or inverse temperature (tDCS: *F*(1,42) = 0.007, *p* = .93, tDCS x Valence: *F*(1,42) = 0.04, *p* = .82)(Figure S15).

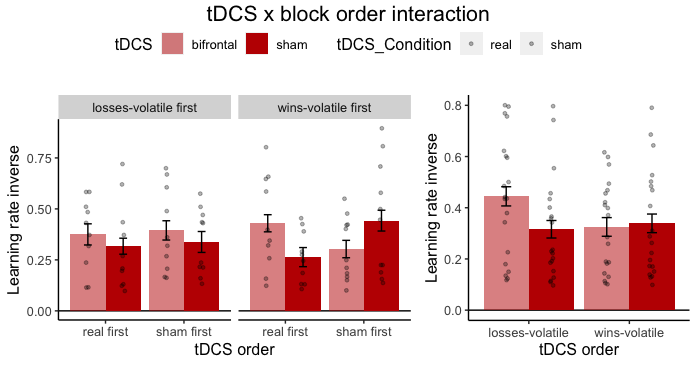

A

C
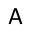

B

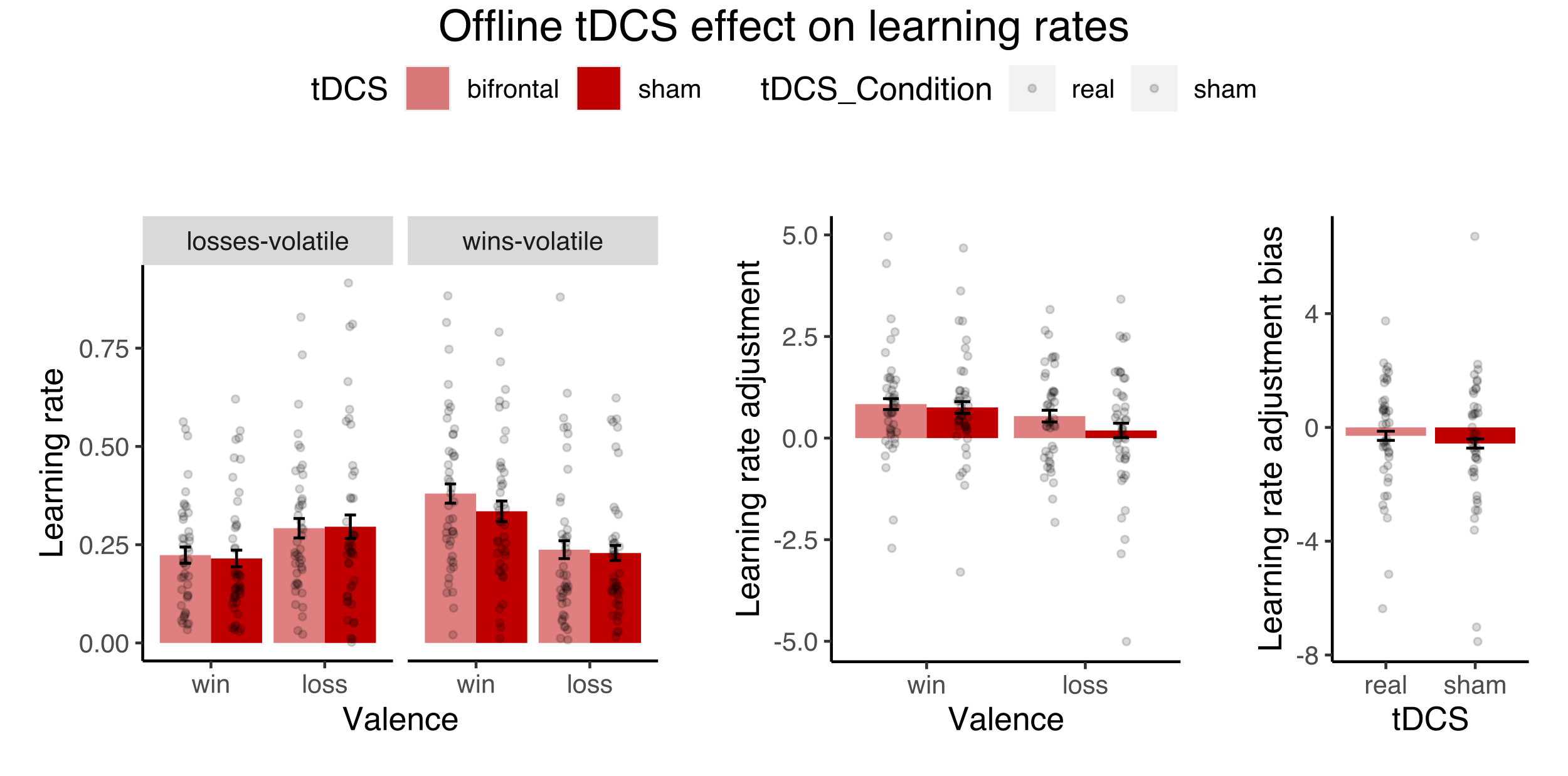

Figure S15. Effects of bifrontal tDCS applied before task performance on learning rates derived from the constant model. Real compared to sham tDCS did not have any significant effect on learning rates (A), learning rate adjustment (B) or learning rate adjustment bias (C).

The key effect of online tDCS was an increase in learning rate adjustment bias, an effect that was not significant in the offline condition. To test directly whether the effect of online bifrontal tDCS was specific to the stimulation time, we conducted a t-test to contrast the effect of real vs. sham tDCS on the learning rate adjustment bias between the online and offline tDCS conditions. The effect of online tDCS was marginally though not significantly larger than the effect of offline tDCS (*t*(78.9) = -1.5, *p* = .063 (Welch two sample t-test, one-sided))(Figure 6, main text).

##### 4.3 The effect of tDCS in the block-wise model

We tested whether the effect of tDCS observed in the constant model was also present in the parameter estimates derived from the block-wise model. To test our main hypothesis that bifrontal tDCS during task performance would increase learning rates from wins, we conducted an ANOVA with tDCS Condition and Valence as factors of interest. There was a significant main effect of tDCS (after removal of five outliers: *F*(1,32) = 5.7, *p* = .022; before outlier removal: *F*(1,37) = 1.5, *p* = .22, see Figure S17) but no interaction between tDCS Condition and Valence (*F*(1,32) = 0.01, *p* = .90). Real compared to sham tDCS increased learning rates independent of valence (Figure S16A).

Figure S16. Effect of online tDCS on learning rates in the block-wise model. (A) Online bifrontal tDCS increased learning rates independent of valence. Online bifrontal tDCS had no effect on learning rate adjustment (B) or learning rate adjustment bias (C).

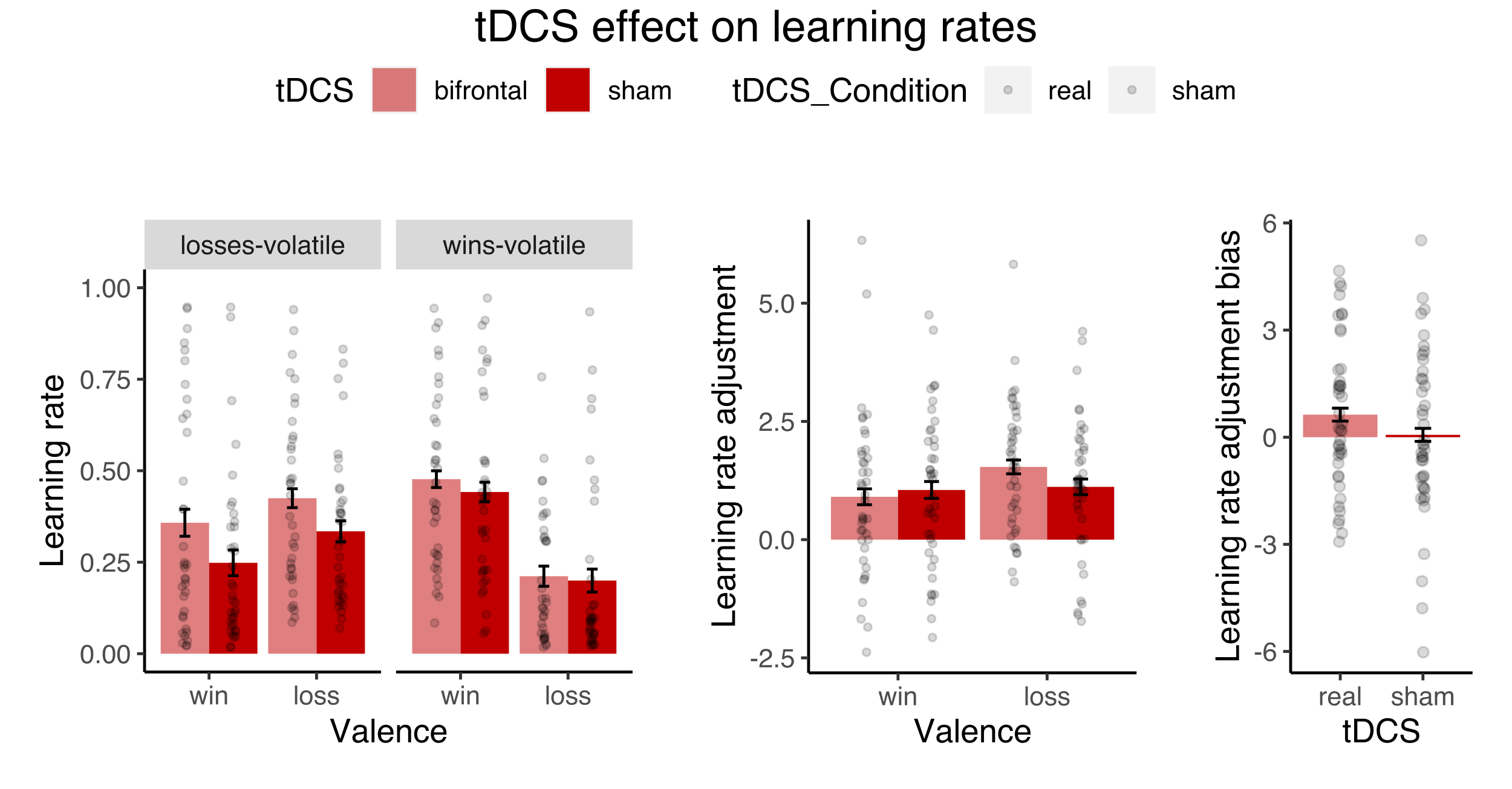

A

C

B

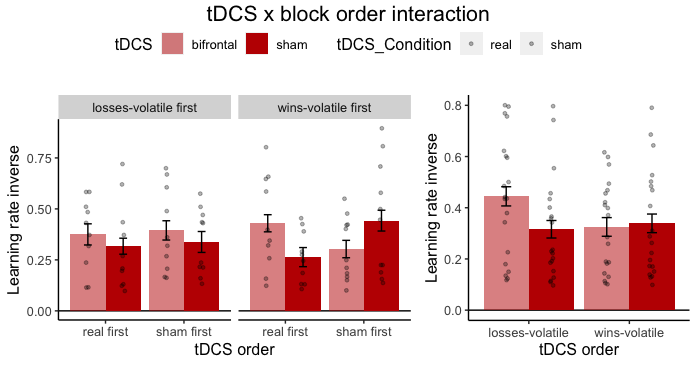

To test whether tDCS might affect learning rate adjustment, we analysed learning rate adjustment as well as the learning rate adjustment bias in separate ANOVAs. There was no main effect of tDCS on learning rate adjustment (*F*(1,37) = 0.36, *p* = .54) nor an interaction between tDCS and Valence (*F*(1,37) = 1.2, *p* = .26)(Figure S16B). There was also no effect of tDCS on the learning rate adjustment bias (*F*(1,37) = 1.2, *p* = .26)(Figure S16C).

We also analysed the inverse temperature to test whether tDCS had an effect on choice randomness. There was no effect of tDCS on the inverse temperature (all *p* > .47).

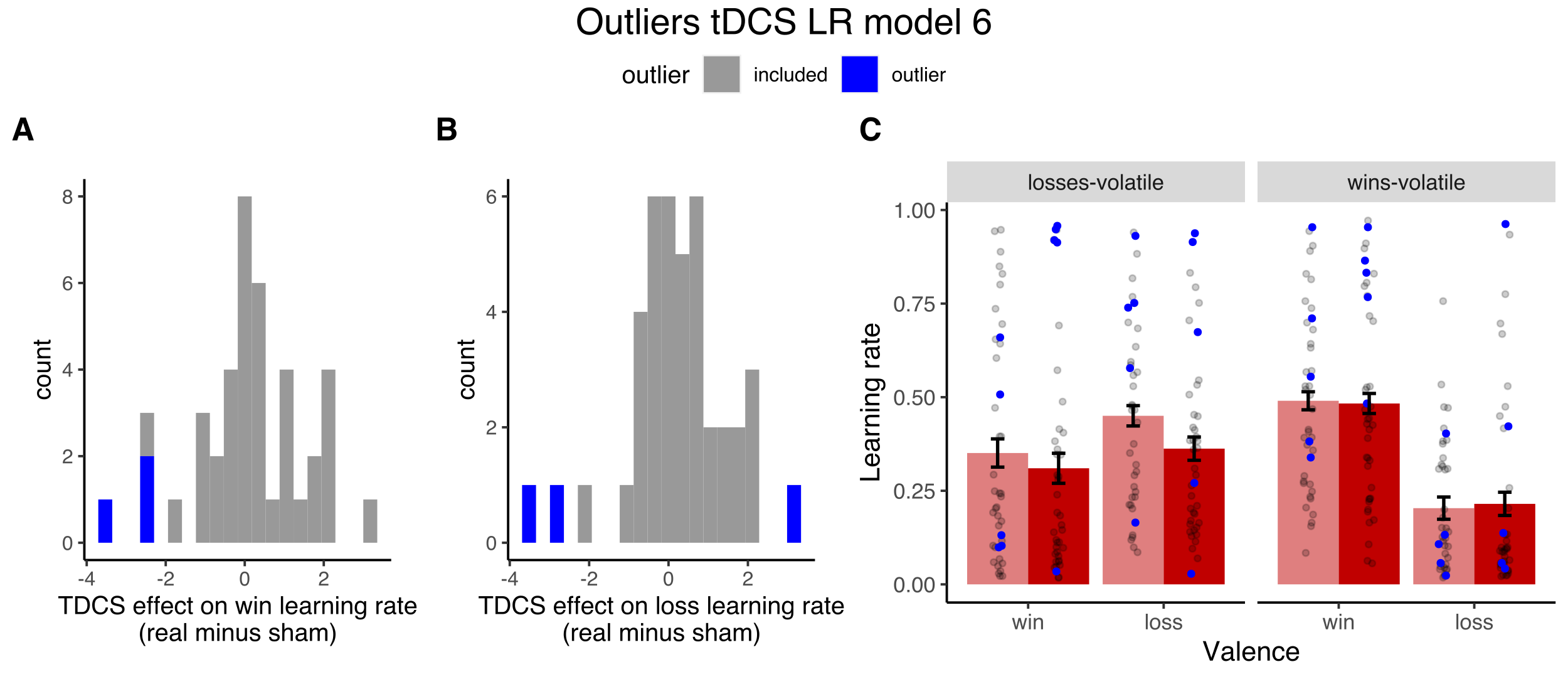

Figure S17. Outlier removal for the effect of tDCS on learning rates derived from the block-wise model (five outliers). Outliers were identified based on the difference in win (A) and loss learning rates (B) between real and sham tDCS. (C) shows the effect of tDCS on learning rates including outliers (see Figure S6.2). Outliers are highlighted in blue. Due to the within-subject design, subjects who were classified as outliers in (A) or (B) were removed entirely from this analysis.

Furthermore, we tested whether tDCS applied before task performance modulated computational measures. By contrast with online tDCS, offline tDCS had no significant effect on learning rates (tDCS: *F*(1,42) = 2.3, *p* = .13, tDCS x Valence: *F*(1,42) = 1.03, *p* = .31, tDCS x Volatility: *F*(1,42) = 0.06, *p* = .79, tDCS x Valence x Volatility: *F*(1,42) = 0.53, *p* = .46), learning rate adjustment (tDCS: *F*(1,42) = 0.5, *p* = .46, tDCS x Valence: *F*(1,42) = 0.06, *p* = .79), learning rate adjustment bias (tDCS: *F*(1,42) = 0.06, *p* = .79) or inverse temperature (tDCS: *F*(1,42) = 0.25, *p* = .61, tDCS x Volatility: *F*(1,42) = 0.03, *p* = .84)(Figure S18).

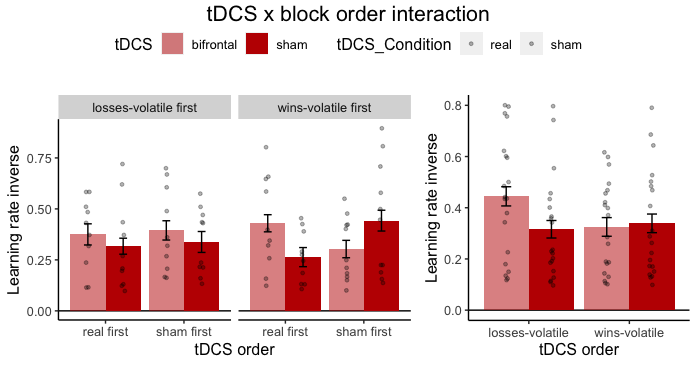

A

B

C

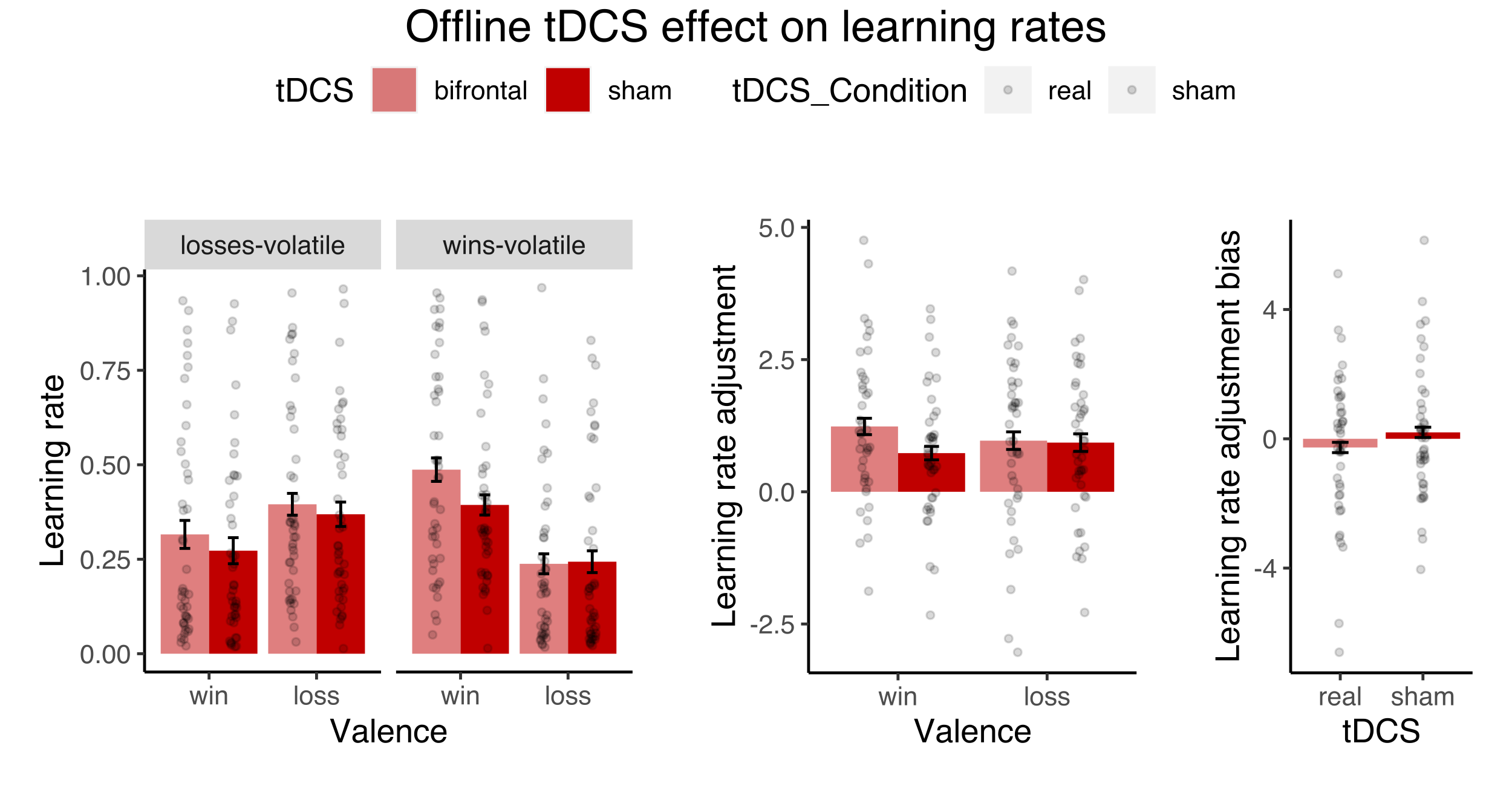

Figure 6.4. Effects of offline bifrontal tDCS on learning rates derived from the block-wise model. Real compared to sham tDCS had no effect on learning rates (A) learning rate adjustment (B) or learning rate adjustment bias (C).

Figure S18. Effects of offline bifrontal tDCS on learning rates derived from the block-wise model. Real compared to sham tDCS had no effect on learning rates (A) learning rate adjustment (B) or learning rate adjustment bias (C).

#### 4.4 The effect of tDCS on mood questionnaires

The state-anxiety scale from the State-Trait Anxiety Inventory (STAI)(5) and Positive and Negative Affect Schedule (PANAS)(6) were completed before and after task performance. Mean scores are included in Table S3. To analyse potential effects of tDCS on mood, we entered the STAI state-anxiety score, the PANAS positive scale and PANAS negative scale into separate ANOVAs including the factors tDCS Condition (real vs. sham) and Time (before vs. after task performance). An effect of tDCS on mood would be indicated by an interaction between Time and tDCS Condition. No significant interaction effect between Time and tDCS Condition were observed for any of the measures for online or offline tDCS (all *p* > .13).

Table S3. Mean (SD) for the STAI state-anxiety scale and PANAS (low mood).

|  | **Online tDCS** | | **Offline tDCS** | |
| --- | --- | --- | --- | --- |
|  | Before | After | Before | After |
| *STAI-S* |  |  |  |  |
| Real | 42.1 (10.0) | 40.1 (8.3) | 41.4 (8.2) | 39.1 (9.1) |
| Sham | 42.1 (11.0) | 38.3 (9.1) | 43.0 (11.6) | 39.2 (10.7) |
| *PANAS positive* |  |  |  |  |
| Real | 24.2 (7.6) | 21.5 (8.0) | 24.2 (6.9) | 23.7 (7.6) |
| Sham | 24.1 (6.8) | 22.3 (7.1) | 23.0 (7.6) | 22 (8.8) |
| *PANAS negative* |  |  |  |  |
| Real | 15.1 (5.9) | 13.3 (4.1) | 15.2 (3.9) | 13.5 (3.9) |
| Sham | 15.1 (5.8) | 13.2 (4.1) | 16.1 (6.1) | 13,2 (4.9) |

STAI-S: State-Trait Anxiety Inventory (State-anxiety scale, range: 20-80); PANAS: Positive and Negative Affect Scale (positive / negative affect scale, range: 10-50)

### 5 Detailed results: Non-computational validation

As non-computational control analyses, logistic regressions were run to predict the choice on each trial using win and loss outcomes of the previous 3 trials:

Choice(n) ~ win(n-1) + loss(n-1) + win(n-2) + loss(n-2) + win(n-3) + loss(n-3)

Regressions were run for each task block and each participant individually. To enable comparison across blocks and participants, the regression weights obtained in each regression were divided by the largest weight obtained in that regression so that weights varied between 0 and 1.

Averaged across all participants, weights for outcomes closer to the choice were larger (i.e. n-1 > n-2 > n-3)(Figure S19). Furthermore, weights depended on the volatility condition. Weights were largest if the respective outcome was volatile and the other stable, and smallest if the respective outcome was stable and the other volatile. A higher learning rate would correspond to a higher weight on the most recent outcome. The following analyses therefore focus mostly on the weights for trial n-1.

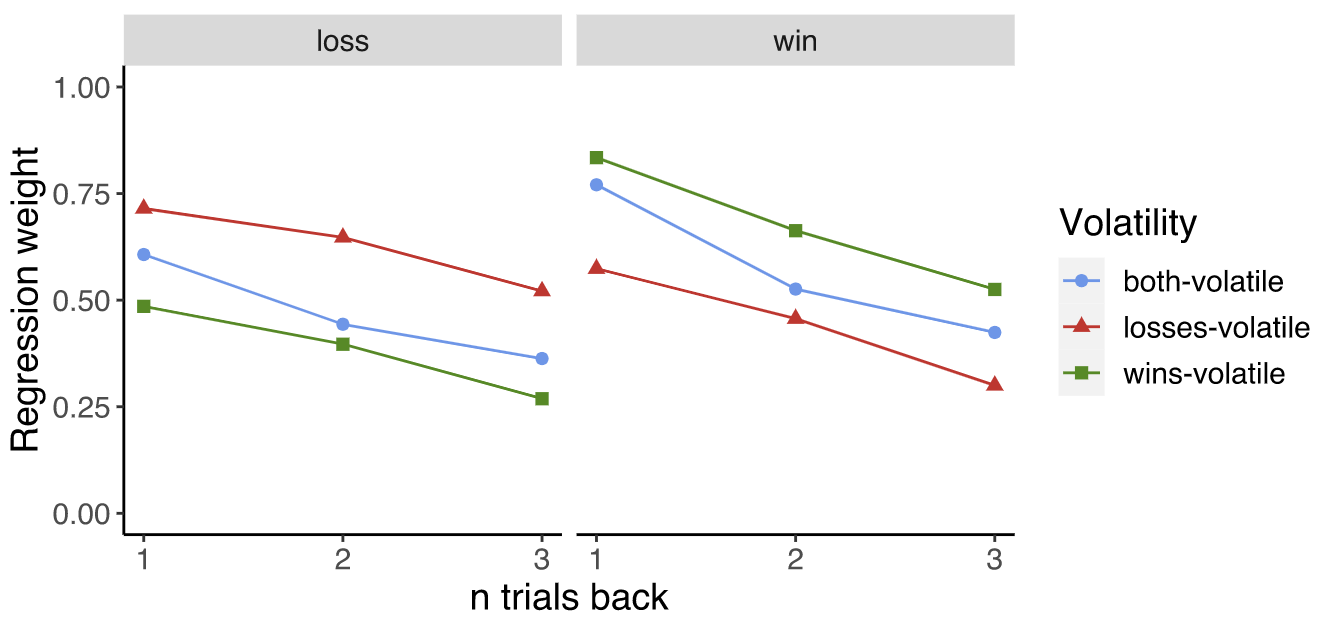

*Figure S19.* Regression weight for the win and loss regressors, 1, 2 and 3 trials back.

#### 5.1 The effect of tDCS on regression weights

In line with the computational analysis, the non-computational analyses focus on the stimulation period (block 2 and 3, wins-volatile and losses-volatile). Weights for all regressors in the real and sham tDCS conditions are shown in Figure S20.

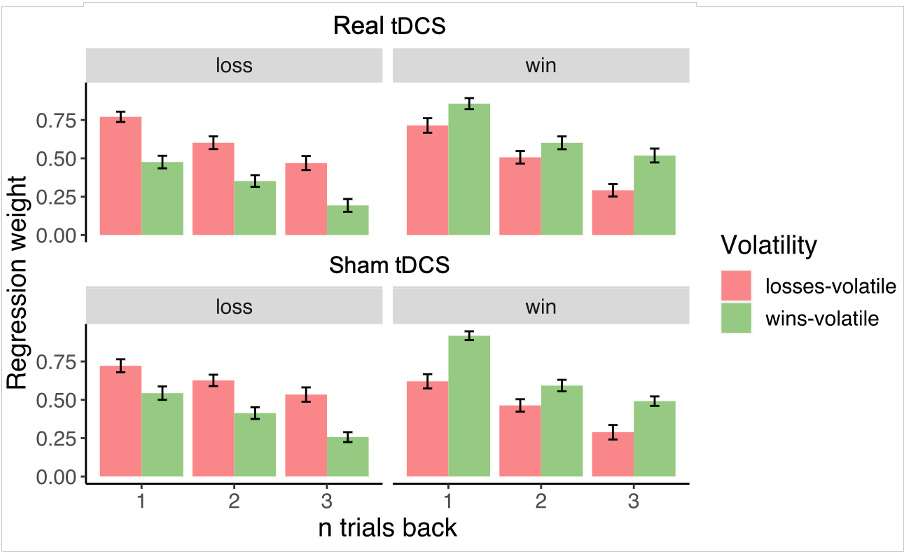

*Figure S20.* Regression weights from the previous three trials during real tDCS (top) and sham tDCS (bottom) for loss outcomes (left) and win outcomes (right).

In the computational analysis, we found that online bifrontal tDCS increased loss learning rate adjustment. As a non-computational equivalent, we defined *weight adjustment* as the difference in the regression weight for trial n-1 for volatile minus stable conditions. For example, *loss weight adjustment* corresponds to the weight on the loss outcome on trial n-1 in the losses-volatile condition, minus the weight on the loss outcome on trial n-1 in the wins-volatile condition.

There was a trend-level effect of tDCS on loss weight adjustment (*t*(37) = 1.87, *p* = .069). Similar to the increase in loss learning rate adjustment, there was a trend towards an increase in loss weight adjustment during real compared to sham tDCS (Figure S21A). The change in weight adjustment for losses induced by tDCS was strongly correlated to the change in loss learning rate adjustment across individuals, indicating that these two measures capture a similar behavioural phenomenon (*r* = .60, *t*(39) = 4.6, *p* < .0001, Figure S21B). In conclusion, the results from the regression analysis support the effect of tDCS on learning rate adjustment observed in the constant model.

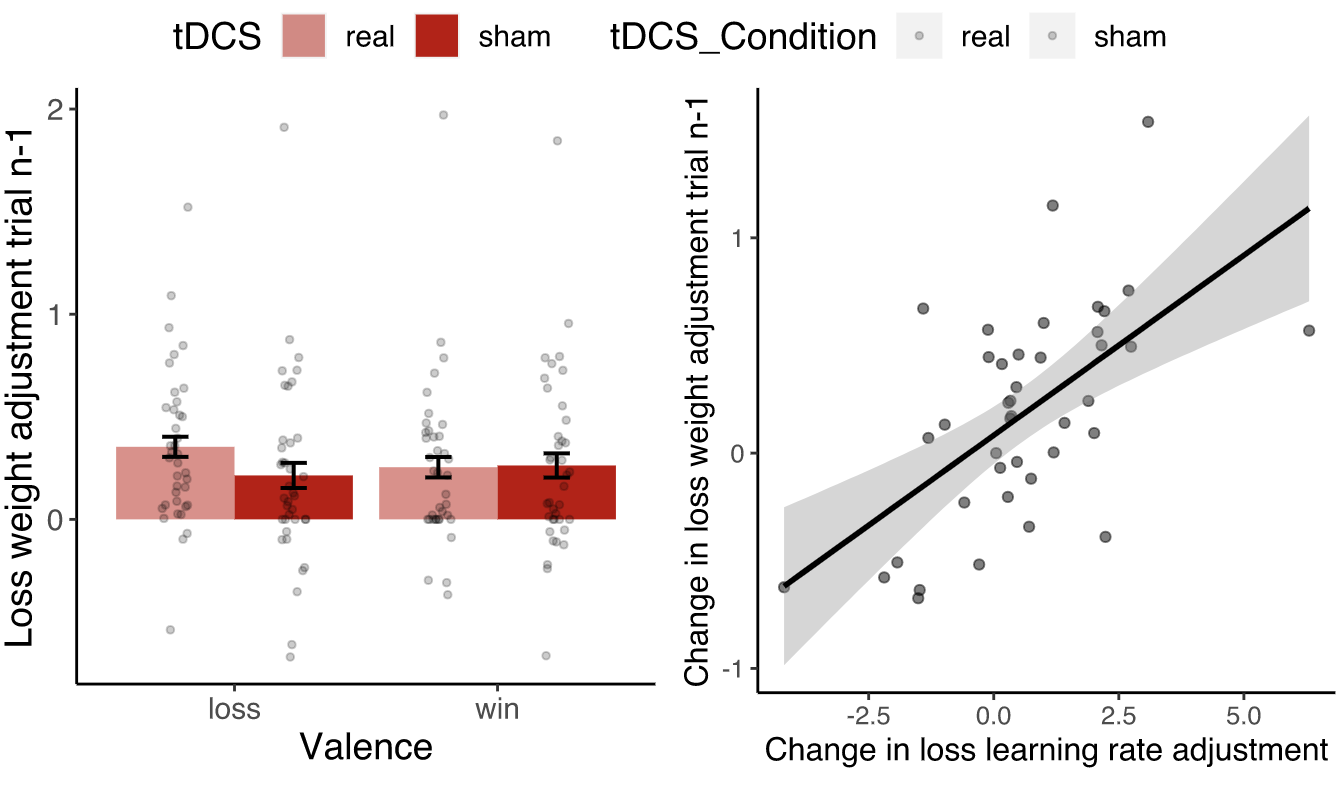

A

B

*p* = .069

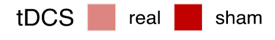

r = .60, *p* < .0001

*Figure S21.* (A) Effect of online bifrontal tDCS on the adjustment of the regression weight for losses on trial n-1. (B) The change in loss weight adjustment correlated significantly with the change in loss learning rate adjustment.

#### 5.2 The effect of low mood on regression weights

Weights for all regressors for the low mood sample and general population sample are shown in Figure S22. In the computational analysis, individuals with low mood showed a lower learning rate adjustment bias than healthy individuals, i.e. lower loss learning rate adjustment and higher win learning rate adjustment. For trial n-1, the low mood sample showed a trend towards lower win weight adjustment than the general population sample (*t*(69,7) = 1.8, *p* = .064, Figure S23A). This is in contrast with the findings in learning rate adjustment.

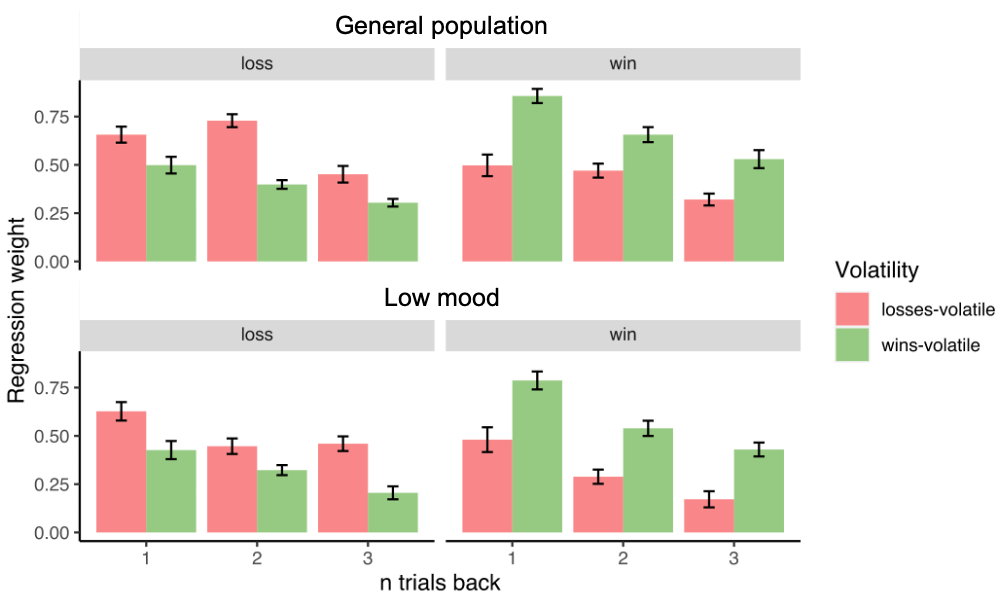

*Figure S22.* Regression weights from the previous three trials for the general population sample (top) and low mood sample (bottom) for loss outcomes (left) and win outcomes (right).

It should be noted that unexpectedly, in the general population the highest weight for loss outcomes was obtained for trial n-2 (not trial n-1 as expected). To test whether the weights for trial n-2 might explain the observed effect on learning rate adjustment, we repeated the same analyses for the regression weight for trial n-2. Here, the low mood sample showed significantly lower loss weight adjustment than the general population sample (*t*(65.5) = 2.6, *p* = .01, Figure S23C) which is in line with the decrease in loss learning rate adjustment observed in the low mood sample. To test whether the group differences in weight adjustment might capture a similar behavioural phenomenon as the differences in learning rate adjustment, we correlated weight adjustment and learning rate adjustment separately for each group. For trial n-1, win weight adjustment was highly correlated with win learning rate adjustment within both samples (general population: r = .84, *t*(38) = 9.7, *p* < .0001; low mood: r = .80, *t*(41) = 8.6, *p* < .0001; Figure S23B). For trial n-2, loss weight adjustment correlated with loss learning rate adjustment only in the general population sample (general population: r = .36, *t*(38) = 2.3, *p* = .022; low mood: r = .21, *t*(41) = 1.4, *p* = .16; Figure S23D). Taken together, the findings from the regression analysis support the observed effect of low mood on learning rate adjustment only in part.

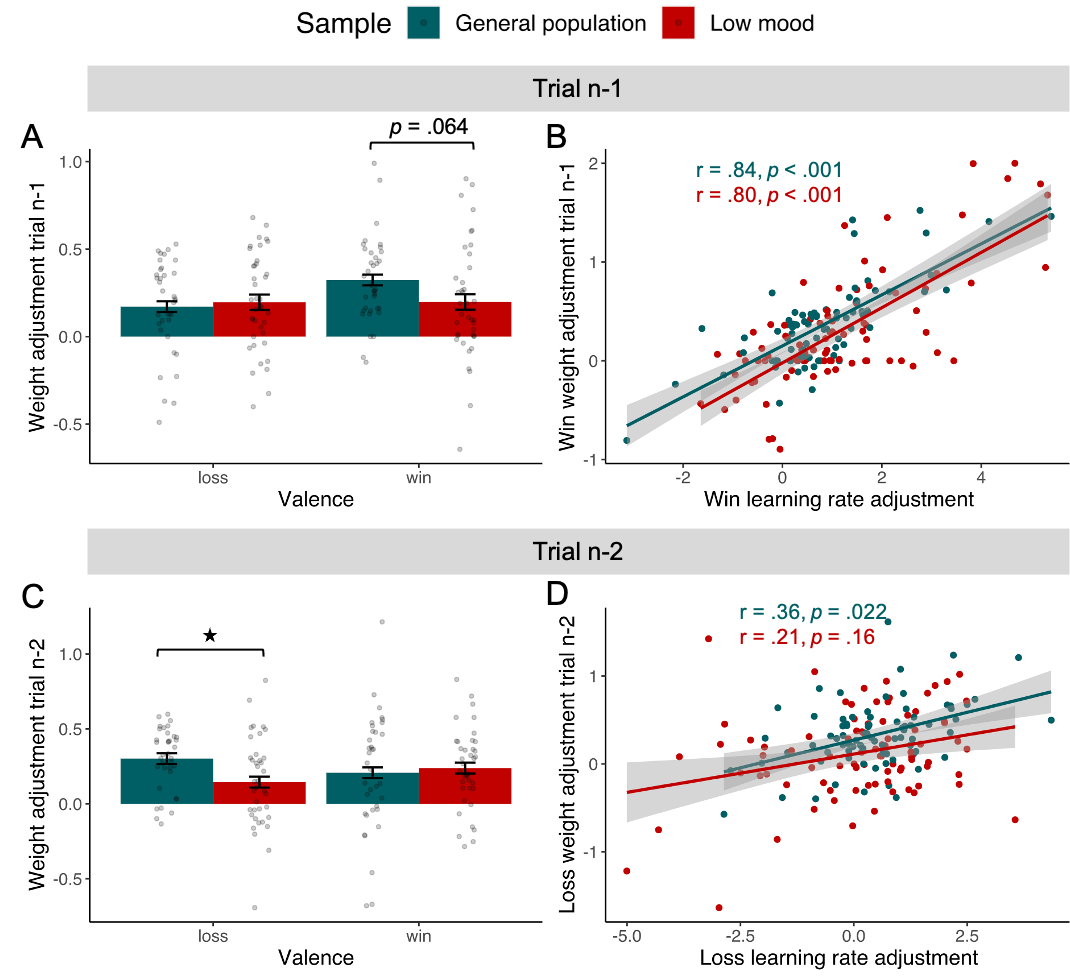

*Figure S23.* Difference in weight adjustment between individuals with low mood and healthy individuals. (A) The low mood sample showed a trend towards lower win weight adjustment (trial n-1) which is in contrast with the observed increase in win learning rate adjustment. (B) Win weight adjustment (n-1) was highly correlated to win learning rate adjustment. (C) For trial n-2, the low mood sample showed significantly lower loss weight adjustment which is in line with the observed decrease in loss learning rate adjustment. (D) Loss weight adjustment for trial n-2 was significantly correlated to loss learning rate adjustment only within the general population sample

### 6 Summary statistics for all computational parameters

#### 6.1 Constant model

Table S4. Mean and standard deviation (SD) for the parameter estimates from the constant model for the comparison between the low mood and general population samples.

| Sample | Volatility | Time | Win LR  mean | Win LR  SD | Loss LR  mean | Loss LR  SD | Win beta mean | Win beta  SD | Loss beta  mean | Loss beta  SD |
| --- | --- | --- | --- | --- | --- | --- | --- | --- | --- | --- |
| General population | both-volatile | 1 | -0.91 | 1.56 | -0.78 | 1.23 | 2.08 | 0.82 | 1.80 | 1.04 |
| General population | both-volatile | 2 | -0.74 | 1.08 | -1.01 | 1.10 | 2.08 | 0.82 | 1.80 | 1.04 |
| General population | losses-volatile | 1 | -1.54 | 1.16 | -1.02 | 0.87 | 2.08 | 0.82 | 1.80 | 1.04 |
| General population | losses-volatile | 2 | -1.71 | 0.99 | -1.03 | 0.82 | 2.08 | 0.82 | 1.80 | 1.04 |
| General population | wins-volatile | 1 | -0.75 | 0.82 | -1.46 | 0.97 | 2.08 | 0.82 | 1.80 | 1.04 |
| General population | wins-volatile | 2 | -0.89 | 0.98 | -1.53 | 1.06 | 2.08 | 0.82 | 1.80 | 1.04 |
| Low mood | both-volatile | 1 | -0.78 | 1.79 | -1.50 | 1.82 | 1.68 | 0.79 | 1.47 | 1.09 |
| Low mood | both-volatile | 2 | -1.12 | 1.43 | -1.07 | 1.34 | 1.68 | 0.79 | 1.47 | 1.09 |
| Low mood | losses-volatile | 1 | -1.59 | 1.25 | -1.19 | 1.70 | 1.68 | 0.79 | 1.47 | 1.09 |
| Low mood | losses-volatile | 2 | -1.79 | 1.29 | -1.54 | 1.21 | 1.68 | 0.79 | 1.47 | 1.09 |
| Low mood | wins-volatile | 1 | -0.49 | 1.22 | -1.29 | 1.17 | 1.68 | 0.79 | 1.47 | 1.09 |
| Low mood | wins-volatile | 2 | -0.55 | 1.22 | -1.70 | 1.02 | 1.68 | 0.79 | 1.47 | 1.09 |

Table S5. Mean and standard deviations for learning rate adjustment (LR adjustment) and learning rate adjustment bias (LR adjustment bias) derived from the constant model for the comparison of the low mood and general population samples.

| Sample | Time | Win LR adjustment  mean | Win LR adjustment  SD | Loss LR adjustment  mean | Loss LR adjustment  SD | LR adjustment bias  mean | LR adjustment bias  SD |
| --- | --- | --- | --- | --- | --- | --- | --- |
| General population | 1 | 0.79 | 1.24 | 0.43 | 1.28 | -0.36 | 1.72 |
| General population | 2 | 0.82 | 1.23 | 0.50 | 1.27 | -0.32 | 1.55 |
| Low mood | 1 | 1.10 | 1.56 | 0.10 | 1.70 | -1.00 | 2.65 |
| Low mood | 2 | 1.24 | 1.54 | 0.16 | 1.61 | -1.08 | 2.22 |

Table S6. Mean and standard deviation (SD) for the parameter estimates from the constant model for the comparison of real vs. sham tDCS applied *during* task performance.

| tDCS Condition | Volatility | Time | Win LR  mean | Win LR  SD | Loss LR  mean | Loss LR  SD | Win beta  mean | Win beta  SD | Loss beta  mean | Loss beta  SD |
| --- | --- | --- | --- | --- | --- | --- | --- | --- | --- | --- |
| real | both-volatile | 1 | -0.52 | 1.11 | -1.20 | 1.65 | 1.99 | 0.73 | 1.69 | 0.98 |
| real | both-volatile | 2 | -0.82 | 1.48 | -1.48 | 1.35 | 1.99 | 0.73 | 1.69 | 0.98 |
| real | losses-volatile | 1 | -1.31 | 1.21 | -0.73 | 0.97 | 1.99 | 0.73 | 1.69 | 0.98 |
| real | losses-volatile | 2 | -1.64 | 1.27 | -0.99 | 1.00 | 1.99 | 0.73 | 1.69 | 0.98 |
| real | wins-volatile | 1 | -0.72 | 1.48 | -1.66 | 1.04 | 1.99 | 0.73 | 1.69 | 0.98 |
| real | wins-volatile | 2 | -0.64 | 1.05 | -1.57 | 0.91 | 1.99 | 0.73 | 1.69 | 0.98 |
| sham | both-volatile | 1 | -0.49 | 1.48 | -1.52 | 1.65 | 1.91 | 0.67 | 1.57 | 1.08 |
| sham | both-volatile | 2 | -0.97 | 1.62 | -1.49 | 1.41 | 1.91 | 0.67 | 1.57 | 1.08 |
| sham | losses-volatile | 1 | -1.47 | 1.33 | -1.05 | 1.11 | 1.91 | 0.67 | 1.57 | 1.08 |
| sham | losses-volatile | 2 | -1.63 | 1.26 | -1.35 | 1.23 | 1.91 | 0.67 | 1.57 | 1.08 |
| sham | wins-volatile | 1 | -0.39 | 1.13 | -1.40 | 1.31 | 1.91 | 0.67 | 1.57 | 1.08 |
| sham | wins-volatile | 2 | -0.69 | 1.42 | -1.66 | 0.98 | 1.91 | 0.67 | 1.57 | 1.08 |

Table S7. Mean and standard deviations for learning rate adjustment (LR adjustment) and learning rate adjustment bias (LR adjustment bias) derived from the constant model for the comparison of real vs. sham tDCS applied *during* task performance.

| tDCS Condition | Time | Win LR adjustment  mean | Win LR adjustment  SD | Loss LR adjustment  mean | Loss LR adjustment  SD | LR adjustment bias  mean | LR adjustment bias  SD |
| --- | --- | --- | --- | --- | --- | --- | --- |
| real | 1 | 0.59 | 1.79 | 0.93 | 1.32 | 0.33 | 2.27 |
| real | 2 | 1.00 | 1.48 | 0.58 | 1.09 | -0.42 | 1.70 |
| sham | 1 | 1.07 | 1.36 | 0.34 | 1.34 | -0.73 | 2.15 |
| sham | 2 | 0.94 | 1.54 | 0.32 | 1.32 | -0.62 | 1.91 |

Table S8. Mean and standard deviation (SD) for the parameter estimates from the constant model for the comparison of real vs. sham tDCS applied *before* task performance.

| tDCS Condition | Volatility | Time | Win LR  mean | Win LR  SD | Loss LR  mean | Loss LR  SD | Win beta  mean | Win beta  SD | Loss beta  mean | Loss beta  SD |
| --- | --- | --- | --- | --- | --- | --- | --- | --- | --- | --- |
| real | both-volatile | 1 | -0.39 | 1.28 | -0.94 | 1.36 | 1.81 | 0.88 | 1.67 | 0.91 |
| real | both-volatile | 2 | -0.68 | 1.22 | -1.16 | 1.42 | 1.81 | 0.88 | 1.67 | 0.91 |
| real | losses-volatile | 1 | -1.45 | 0.90 | -1.05 | 1.01 | 1.81 | 0.88 | 1.67 | 0.91 |
| real | losses-volatile | 2 | -1.69 | 1.35 | -0.86 | 0.96 | 1.81 | 0.88 | 1.67 | 0.91 |
| real | wins-volatile | 1 | -0.59 | 1.04 | -1.50 | 1.30 | 1.81 | 0.88 | 1.67 | 0.91 |
| real | wins-volatile | 2 | -0.75 | 1.15 | -1.39 | 1.11 | 1.81 | 0.88 | 1.67 | 0.91 |
| sham | both-volatile | 1 | -0.63 | 1.47 | -0.88 | 1.46 | 1.80 | 0.96 | 1.70 | 1.06 |
| sham | both-volatile | 2 | -1.14 | 1.06 | -1.27 | 1.37 | 1.80 | 0.96 | 1.70 | 1.06 |
| sham | losses-volatile | 1 | -1.55 | 1.02 | -1.20 | 1.56 | 1.80 | 0.96 | 1.70 | 1.06 |
| sham | losses-volatile | 2 | -1.77 | 1.07 | -1.18 | 0.96 | 1.80 | 0.96 | 1.70 | 1.06 |
| sham | wins-volatile | 1 | -0.85 | 1.07 | -1.47 | 1.05 | 1.80 | 0.96 | 1.70 | 1.06 |
| sham | wins-volatile | 2 | -0.80 | 0.89 | -1.67 | 1.10 | 1.80 | 0.96 | 1.70 | 1.06 |

Table S9. Mean and standard deviations for learning rate adjustment (LR adjustment) and learning rate adjustment bias (LR adjustment bias) derived from the constant model for the comparison of real vs. sham tDCS applied *before* task performance.

| tDCS Condition | Time | Win LR adjustment  mean | Win LR adjustment  SD | Loss LR adjustment  mean | Loss LR adjustment  SD | LR adjustment bias  mean | LR adjustment bias  SD |
| --- | --- | --- | --- | --- | --- | --- | --- |
| real | 1 | 0.86 | 1.34 | 0.44 | 1.33 | -0.42 | 2.11 |
| real | 2 | 0.94 | 1.54 | 0.53 | 0.90 | -0.41 | 1.97 |
| sham | 1 | 0.70 | 1.37 | 0.26 | 1.62 | -0.44 | 2.52 |
| sham | 2 | 0.97 | 1.37 | 0.49 | 1.49 | -0.49 | 2.33 |

#### 6.2 Block-wise model

Table S10. Mean and standard deviation (SD) for the parameter estimates from the block-wise model for the comparison between the low mood and general population samples.

| Sample | Volatility | Time | Win LR  mean | Win LR  SD | | Loss LR  mean | Loss LR SD | Beta  mean | Beta  SD | Tendency mean | Tendency SD |
| --- | --- | --- | --- | --- | --- | --- | --- | --- | --- | --- | --- |
| General population | both-volatile | 1 | -0.34 | | 1.78 | -0.66 | 1.52 | 2.19 | 1.15 | -0.03 | 0.13 |
| General population | both-volatile | 2 | -0.24 | | 1.37 | -1.03 | 1.51 | 2.28 | 1.36 | 0.03 | 0.15 |
| General population | losses-volatile | 1 | -1.18 | | 1.35 | -0.69 | 1.01 | 1.56 | 1.72 | 0.05 | 0.17 |
| General population | losses-volatile | 2 | -1.44 | | 1.41 | -0.74 | 1.07 | 1.88 | 1.72 | -0.01 | 0.21 |
| General population | wins-volatile | 1 | -0.45 | | 1.17 | -1.93 | 1.33 | 2.18 | 1.33 | 0.03 | 0.13 |
| General population | wins-volatile | 2 | -0.45 | | 1.33 | -1.90 | 1.48 | 2.00 | 1.35 | -0.04 | 0.18 |
| Low mood | both-volatile | 1 | 0.32 | | 1.70 | -0.65 | 1.86 | 1.37 | 1.17 | -0.05 | 0.19 |
| Low mood | both-volatile | 2 | -0.39 | | 1.42 | -0.70 | 1.57 | 1.76 | 1.47 | 0.02 | 0.16 |
| Low mood | losses-volatile | 1 | -1.07 | | 1.76 | -0.33 | 1.44 | 1.33 | 1.62 | -0.01 | 0.20 |
| Low mood | losses-volatile | 2 | -1.20 | | 1.73 | -0.72 | 0.93 | 1.22 | 1.90 | 0.06 | 0.21 |
| Low mood | wins-volatile | 1 | 0.07 | | 1.47 | -1.38 | 1.65 | 1.66 | 1.34 | -0.04 | 0.18 |
| Low mood | wins-volatile | 2 | -0.08 | | 1.71 | -1.90 | 1.60 | 1.72 | 1.42 | 0.01 | 0.13 |

Table S11. Mean and standard deviations for learning rate adjustment (LR adjustment) and learning rate adjustment bias (LR adjustment bias) derived from the block-wise model for the comparison of the low mood and general population samples.

| Sample | Time | Win LR adjustment  mean | Win LR adjustment  SD | Loss LR adjustment  mean | Loss LR adjustment SD | LR adjustment bias  mean | LR adjustment bias  SD |
| --- | --- | --- | --- | --- | --- | --- | --- |
| General population | 1 | 0.74 | 1.30 | 1.24 | 1.58 | 0.51 | 1.78 |
| General population | 2 | 0.99 | 1.33 | 1.17 | 1.32 | 0.18 | 1.50 |
| Low mood | 1 | 1.14 | 1.73 | 1.05 | 1.56 | -0.09 | 2.52 |
| Low mood | 2 | 1.12 | 1.83 | 1.18 | 1.59 | 0.06 | 2.49 |

Table S12. Mean and standard deviation (SD) for the parameter estimates from the block-wise model for the comparison of real vs. sham tDCS applied *during* task performance.

| tDCS Condition | Volatility | Time | Win LR  mean | Win LR  SD | Loss LR  mean | Loss LR  SD | Beta  mean | Beta  SD | Tendency mean | Tendency SD |
| --- | --- | --- | --- | --- | --- | --- | --- | --- | --- | --- |
| real | both-volatile | 1 | -0.01 | 1.34 | -1.09 | 1.74 | 2.10 | 1.19 | -0.01 | 0.15 |
| real | both-volatile | 2 | -0.50 | 1.66 | -1.70 | 1.62 | 2.22 | 1.20 | 0.02 | 0.15 |
| real | losses-volatile | 1 | -0.90 | 1.79 | -0.23 | 1.29 | 1.66 | 1.58 | 0.05 | 0.20 |
| real | losses-volatile | 2 | -1.28 | 1.62 | -0.71 | 1.44 | 1.74 | 1.55 | 0.01 | 0.17 |
| real | wins-volatile | 1 | 0.01 | 1.24 | -1.77 | 1.26 | 1.83 | 1.54 | -0.01 | 0.15 |
| real | wins-volatile | 2 | -0.34 | 1.39 | -2.06 | 1.30 | 2.07 | 1.15 | 0.00 | 0.14 |
| sham | both-volatile | 1 | 0.39 | 1.57 | -1.27 | 1.70 | 1.84 | 1.24 | -0.05 | 0.14 |
| sham | both-volatile | 2 | -0.35 | 1.54 | -1.50 | 1.66 | 1.94 | 1.24 | 0.01 | 0.16 |
| sham | losses-volatile | 1 | -1.08 | 1.92 | -0.68 | 1.31 | 1.71 | 1.33 | -0.02 | 0.15 |
| sham | losses-volatile | 2 | -1.16 | 1.78 | -0.85 | 1.08 | 1.49 | 1.74 | 0.09 | 0.16 |
| sham | wins-volatile | 1 | -0.03 | 1.48 | -1.79 | 1.71 | 2.00 | 1.06 | 0.02 | 0.13 |
| sham | wins-volatile | 2 | -0.14 | 1.85 | -2.01 | 1.43 | 1.99 | 1.28 | 0.04 | 0.17 |

Table S13. Mean and standard deviations for learning rate adjustment (LR adjustment) and learning rate adjustment bias (LR adjustment bias) derived from the block-wise model for the comparison of real vs. sham tDCS applied *during* task performance.

| tDCS Condition | Time | Win LR adjustment mean | Win LR adjustment  SD | Loss LR adjustment  mean | Loss LR adjustment  SD | LR adjustment bias  mean | LR adjustment bias  SD |
| --- | --- | --- | --- | --- | --- | --- | --- |
| real | 1 | 0.91 | 1.71 | 1.54 | 1.38 | 0.63 | 2.10 |
| real | 2 | 0.94 | 1.52 | 1.35 | 1.66 | 0.40 | 2.07 |
| sham | 1 | 1.05 | 1.65 | 1.12 | 1.47 | 0.07 | 2.37 |
| sham | 2 | 1.02 | 1.92 | 1.15 | 1.44 | 0.13 | 2.43 |

Table S14. Mean and standard deviation (SD) for the parameter estimates from the block-wise model for the comparison of real vs. sham tDCS applied *before* task performance.

| tDCS Condition | Volatility | Time | Win LR  mean | Win LR  SD | Loss LR mean | Loss LR  SD | Beta  mean | Beta  SD | Tendency  mean | Tendency SD |
| --- | --- | --- | --- | --- | --- | --- | --- | --- | --- | --- |
| real | both-volatile | 1 | -0.01 | 1.71 | -0.97 | 1.77 | 1.95 | 1.03 | -0.03 | 0.19 |
| real | both-volatile | 2 | -0.41 | 1.52 | -0.93 | 1.67 | 2.19 | 1.22 | -0.02 | 0.14 |
| real | losses-volatile | 1 | -1.10 | 1.65 | -0.51 | 1.35 | 1.55 | 1.58 | 0.09 | 0.21 |
| real | losses-volatile | 2 | -1.49 | 1.75 | -0.45 | 1.51 | 1.82 | 1.65 | -0.01 | 0.13 |
| real | wins-volatile | 1 | -0.02 | 1.43 | -1.53 | 1.48 | 1.78 | 1.36 | 0.00 | 0.20 |
| real | wins-volatile | 2 | -0.35 | 1.37 | -1.53 | 1.69 | 1.80 | 1.45 | -0.03 | 0.18 |
| sham | both-volatile | 1 | -0.17 | 1.69 | -0.67 | 1.74 | 1.93 | 1.25 | -0.01 | 0.17 |
| sham | both-volatile | 2 | -0.62 | 1.29 | -0.92 | 1.74 | 1.93 | 1.45 | -0.01 | 0.14 |
| sham | losses-volatile | 1 | -1.35 | 1.56 | -0.67 | 1.38 | 1.68 | 1.68 | 0.04 | 0.21 |
| sham | losses-volatile | 2 | -1.62 | 1.55 | -0.83 | 1.16 | 1.74 | 1.82 | 0.03 | 0.18 |
| sham | wins-volatile | 1 | -0.49 | 1.24 | -1.61 | 1.52 | 1.86 | 1.43 | -0.06 | 0.15 |
| sham | wins-volatile | 2 | -0.43 | 1.18 | -1.80 | 1.66 | 1.88 | 1.45 | -0.01 | 0.14 |

Table S15. Mean and standard deviations for learning rate adjustment (LR adjustment) and learning rate adjustment bias (LR adjustment bias) derived from the block-wise model for the comparison of real vs. sham tDCS applied *before* task performance.

| tDCS Condition | Time | Win LR adjustment  mean | Win LR adjustment  SD | Loss LR adjustment  mean | Loss LR adjustment  SD | LR adjustment bias  mean | LR adjustment bias  SD |
| --- | --- | --- | --- | --- | --- | --- | --- |
| real | 1 | 1.07 | 1.57 | 1.02 | 1.59 | -0.05 | 2.45 |
| real | 2 | 1.14 | 1.39 | 1.08 | 1.38 | -0.06 | 1.89 |
| sham | 1 | 0.87 | 1.45 | 0.95 | 1.37 | 0.08 | 2.16 |
| sham | 2 | 1.19 | 1.50 | 0.97 | 1.67 | -0.22 | 2.64 |
